# SingleBrain: A Meta-Analysis of Single-Nucleus eQTLs Linking Genetic Risk to Brain Disorders

**DOI:** 10.1101/2025.03.06.25323424

**Authors:** Beomjin Jang, BP Kailash, Alex Tokolyi, Winston H. Cuddleston, Ashvin Ravi, Sang-Hyuk Jung, Tatsuhiko Naito, Beomsu Kim, Min Seo Kim, Minyoung Cho, Mi-So Park, Mikaela Rosen, Joel Blanchard, Jack Humphrey, David A Knowles, Hong-Hee Won, Towfique Raj

## Abstract

Most genetic risk variants for neurological diseases are located in non-coding regulatory regions, where they may often act as expression quantitative trait loci (eQTLs), modulating gene expression and influencing disease susceptibility. However, eQTL studies in bulk brain tissue or specific cell types lack the resolution to capture the brain’s cellular diversity. Single-nucleus RNA sequencing (snRNA-seq) offers high-resolution mapping of eQTLs across diverse brain cell types. Here, we performed a meta-analysis, “SingleBrain,” integrating publicly available snRNA-seq and genotype data from four cohorts, totaling 5.8 million nuclei from 983 individuals. We mapped *cis*-eQTLs across major brain cell types and subtypes and employed statistical colocalization and Mendelian randomization to identify genes mediating neurological disease risk. We observed up to a 10-fold increase in *cis*-eQTLs compared to previous studies and uncovered novel cell type-specific genes linked to Alzheimer’s disease, Parkinson’s disease, and schizophrenia that were previously undetectable in bulk tissue analyses. Additionally, we prioritized putative causal variants for each locus through fine-mapping and integration with cell type-specific enhancer and promoter regulatory elements. SingleBrain represents a comprehensive single-cell eQTL resource, advancing insights into the genetic regulation of brain disorders.

## Introduction

Genome-wide association studies (GWAS) have identified hundreds of genetic risk variants associated with neurological diseases, the majority of which are located in non-coding regions of the genome. These non-coding variants can exert their effects by regulating gene expression through expression quantitative trait loci (eQTLs). eQTLs provide a critical link between non-coding variants and their functional impact on cellular processes. While eQTL studies using bulk brain tissues^1–4^ and specific brain cell types such as primary human microglia^5–8^ or induced pluripotent stem cell (iPSC)-derived models^9,10^ have offered valuable insights into the regulatory mechanisms underlying disease, these approaches are often limited in their representation of the brain’s diverse cell types.

Single-nucleus RNA-seq (snRNA-seq) eQTL studies enable the direct association of genetic variants with gene expression at single-cell-type resolution in the human brain. These studies leverage cell-type level data to uncover the molecular mechanisms of disease-associated variants and enhance our understanding of the cellular and molecular pathways implicated in complex brain disorders. Although recent snRNA-seq eQTL studies have identified hundreds of eQTLs in postmortem human brains^11–13^, larger sample sizes and increased numbers of nuclei are essential to improve statistical power and uncover cell type-specific eQTL effects, particularly in rarer cell populations.

In this study, we conducted a comprehensive *cis*-eQTL meta-analysis, referred to as “SingleBrain”, by integrating publicly available snRNA-seq and genotype data from four independent cohorts: *Fujita* et al.^11^, *Mathys* et al.^14^, *Gabitto* et al.^15^, and *Bryois* et al.^12^ (**Figure 1**). This dataset included 5.8 million nuclei from 983 donors of European ancestry. Using linear mixed models^16^, we performed eQTL analyses across seven major neocortical cell types and 28 subtypes. We subsequently applied colocalization, Mendelian randomization (MR), and fine-mapping analyses to prioritize genes and genetic variants that influence susceptibility to neurodegenerative and neuropsychiatric diseases through their effects on gene expression.

**Figure 1.**
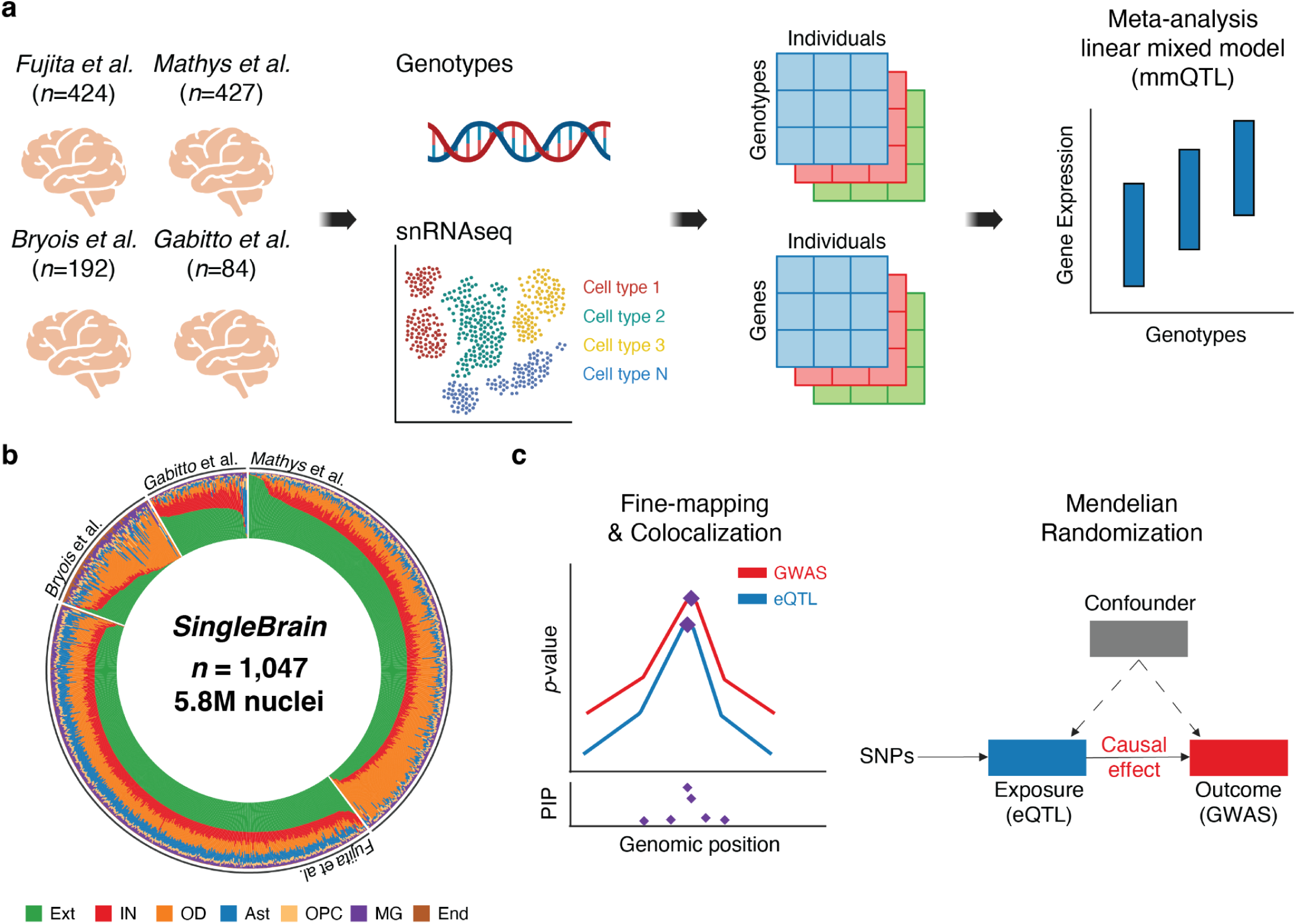
Overview of SingleBrain. **a,** Integration of publicly available single-nucleus RNA sequencing (snRNA-seq) and genotype data from four independent studies^11,12,14,15^. A *cis*-eQTL meta-analysis was conducted across major central nervous system (CNS) cells and subtypes using a linear mixed model approach (multivariate multiple QTL, mmQTL^16^). **b,** Cell proportion of each donor. **c,** Candidate genes and putative causal variants associated with neurodegenerative and neuropsychiatric diseases were identified through statistical colocalization, Mendelian randomization, and fine-mapping. Ext, excitatory neurons; IN, inhibitory neurons; OD, oligodendrocytes; Ast, astrocytes; OPC, oligodendrocyte progenitor cells; MG, microglia; End, endothelial cells.

**SingleBrain** represents the most comprehensive resource to date for understanding cell-type specific *cis*-regulatory effects in the human brain transcriptome. It serves as an important tool for advancing mechanistic understanding of neurological diseases.

## Results

### Single-nucleus expression quantitative loci meta-analysis across multiple brain cohorts

To construct the **‘SingleBrain’** resource, we integrated snRNA-seq data and genotypes from four published datasets comprising 1,047 brain samples from 983 individuals (**Figure 1a and Supplementary Table 1**). The datasets include *Fujita* et al.^11^ and *Mathys* et al.^14^, which used samples from the Religious Orders Study and Memory and Aging Project (ROSMAP), as well as *Bryois* et al.^12^ and *Gabitto* et al.^15^. All snRNA-seq data were generated from frozen samples of the dorsolateral prefrontal cortex (DLPFC) or middle temporal gyrus (MTG) regions. Across the four studies, 207 individuals had mild cognitive impairment (MCI), 336 had a pathological diagnosis of Alzheimer’s disease (AD), 342 were cognitively normal, 33 other dementia and 65 were unknown. 579 individuals were female (**Supplementary Figure 1 and Supplementary Table 2**). Following preprocessing and stringent quality control (QC), 983 participants with both snRNA-seq data and either single-nucleotide polymorphism (SNP) array genotyping (*Gabitto* et al. and *Bryois* et al.) or whole-genome sequencing (WGS) were retained for downstream analyses. *Fujita* et al. and *Mathys* et al. cohorts shared 226 overlapping donors, which were accounted for in the linear mixed model^16^. Following genotype imputation for the array chip dataset, we obtained 5.9M–8.6M SNPs with a minor allele frequency greater than 0.01 for each dataset (**Supplementary Table 1**). Only samples of European ancestry were included (**Supplementary Figure 2 and Supplementary Table 2**). For the snRNA-seq data, QC processes—including demultiplexing, doublet removal, and filtering—resulted in 5.8 million nuclei spanning seven major brain cell types (**Figure 1b and Supplementary Table 3**). To identify candidate genes and putative causal variants associated with neurodegenerative and neuropsychiatric diseases, we performed statistical colocalization, MR, and fine-mapping analyses (**Figure 1c**).

### Cell-type specific eQTL mapping in the human brain

We performed *cis*-eQTL analysis using pseudobulk RNA expression by aggregating unique molecular identifier (UMI) counts per individual per gene per major cell type (or subtype) (see Methods). A linear mixed model^16^ was used to perform meta-analysis across the four datasets. The number of eQTL genes (eGenes) with statistically significant eQTLs at false discovery rate (FDR) < 0.05 ranged from 11,629 in excitatory neurons (Ext) to 4,258 in endothelial cells (End) (**Figures 2a, 2c, Supplementary Figure 3 and Supplementary Table 4**). As expected, the number of significant eGenes increases with greater cell type abundance (**Figure 2a**). SingleBrain demonstrates a 3.21- to 10.42-fold increase in eQTL discovery compared to previous studies^11,12^, highlighting the impact of an expanded sample size (**Figure 2b**).

**Figure 2.**
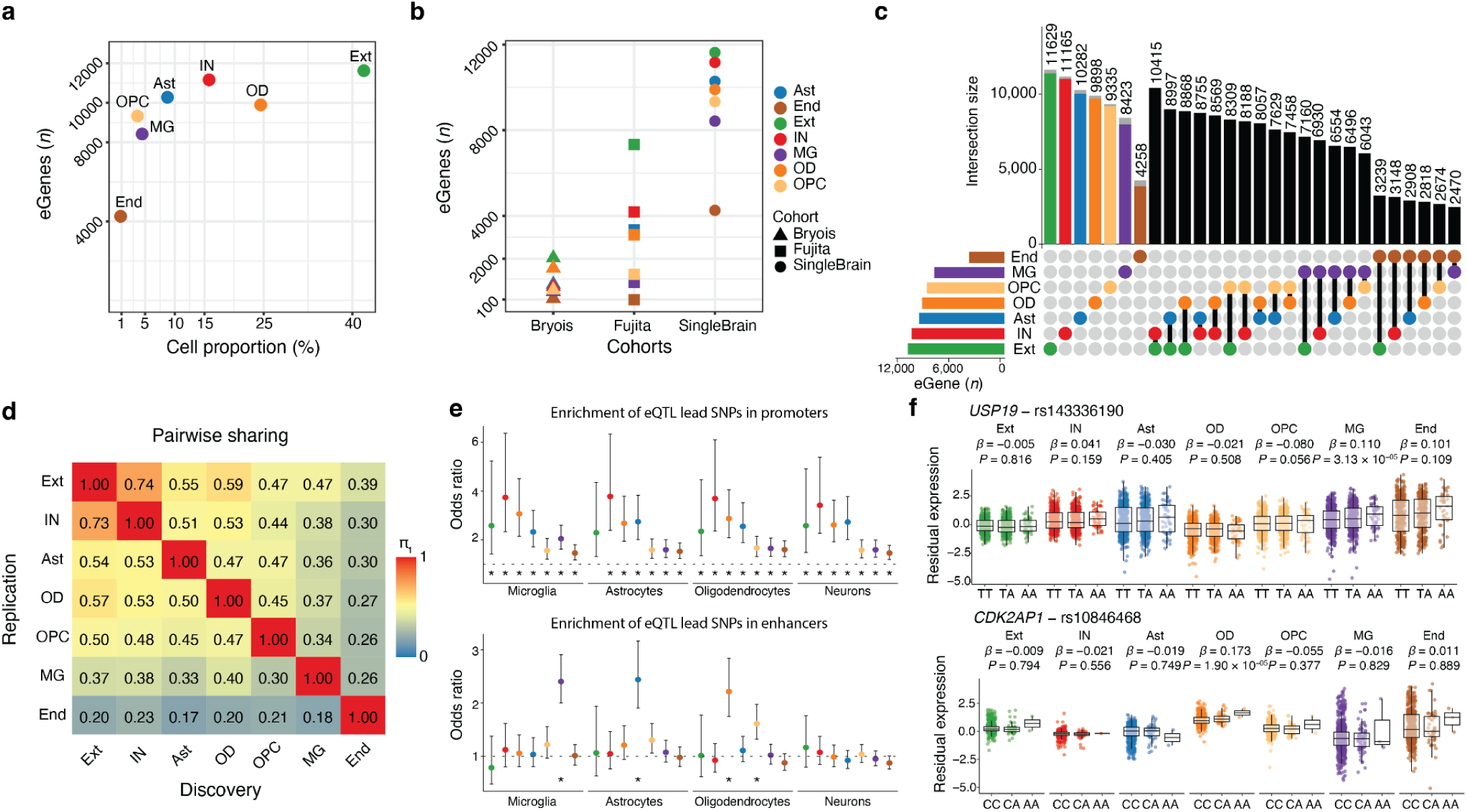
SingleBrain *cis*-eQTLs. **a**, Number of eGenes identified as a function of cell type proportion (%). **b**, Comparison of the number of eGenes (q-value <0.05) identified in this study (SingleBrain), *Bryois* et al., and *Fujita* et al.. **c**, Number of eGenes identified per cell type and shared across cell types. The left bar plot is the number of eGenes by each cell type. The gray color of the barplot presents cell type-specific eGenes within the major cell type. **d**, Pairwise sharing of genes with significant *cis*-eQTLs across major cell types. The numbers shown are Storey’s π_1_ for each cell-type pair. π_1_ is an estimate of the proportion of true alternative hypotheses in the replication cell-type, derived from the distribution of *P*-values. The columns are used for brain major eQTL lead SNPs of this study, and the replication rate is each major cell type QTLs in discovery SNPs, respectively. **e**, Enrichment of cell type-specific eQTL lead SNPs in promoters (top) and enhancers (bottom) of brain cell types, as identified by *Nott et al.*^17^ The dashed line indicates an odds ratio of 1. Enrichment was assessed using Fisher’s Exact Test, with two-sided *P-values* Bonferroni-corrected. Bars indicate 95% confidence intervals of the odds ratio. **f,** Examples of cell-type-specific *cis*-eQTLs. The residual expression is PEER adjusted gene expression level. The nominal *P*-value and beta from the linear regression model in the brain cell type eQTL analysis are indicated above the box plots. The box plots show the median, the box spans from the first to the third quartiles, and the whiskers extend 1.5 times the interquartile range (IQR) from the box. Ext, excitatory neurons; IN, inhibitory neurons; OD, oligodendrocytes; Ast, astrocytes; OPC, oligodendrocyte progenitor cells; MG, microglia; End, endothelial cells.

To characterize the regulatory landscape of eQTLs, we examined the genomic distribution of lead eQTL SNPs and their enrichment in functional elements. The lead eQTL SNPs are predominantly located near the transcription start sites of their target genes^18,19^ (**Supplementary Figure 4**). Furthermore, using cell-type-specific epigenomic data^17^, we find lead eQTL SNPs associated with cell-type-specific eQTLs exhibit significant enrichment within their respective promoters and enhancers (**Figure 2e and Supplementary Table 5**).

We observed a substantial overlap in the eGenes discovered in SingleBrain with previously published eQTL studies^11,12^ (π_1_ = 0.81-0.97), supporting the robustness and reproducibility of our findings (**Supplementary Figure 5)**. A high degree of sharing was observed between some of the major cell types in SingleBrain (Storey’s π_1_ = 0.17-0.74) and subtypes within each major cell type (Storey’s π_1_ = 0.11-0.89) (**Figure 2d and Supplementary Figure 6**). The strongest overlap was detected between excitatory neurons and inhibitory neurons (Storey’s π_1_ = 0.74). After excluding endothelial cells, the lowest sharing (π_1_ = 0.30) was observed between oligodendrocyte progenitor cells and microglia. Endothelial cells exhibited the least sharing across all cell types, likely at least partially due to limited statistical power. We identified a total of 1,793 cell type-specific eQTLs, which are eQTLs detected in one only cell type (**Figure 2f**).

In addition to analyzing major cell types, we performed eQTL analysis across 28 distinct cellular subtypes using the subtype annotations from *Gabitto* et al.^15^ and *Green* et al.^20^. The summary of *cis*-eQTLs identified in each of the 28 subtypes is listed in **Supplementary Table 6**. At the subtype level, 11,118 eGenes were detected in excitatory neurons 1 (Ext1,layer 2/3 intratelencephalic neurons)^15^ but only 6,863 eGenes in Ext6 (layer 6 corticothalamic)^15^. The identification of eQTLs in specific subtypes is strongly influenced by cell type abundance, limiting our power to detect eQTLs in some subtypes (**Supplementary Figure 3 and Supplementary Table 6**). Therefore, we only had the power to identify *cis*-eQTLs for four microglia subtypes (MG1, surveilling; MG2, reacting; MG3, enhanced redox; and MG4, lipid processing; defined by *Green* et al.^20^) (**Supplementary Figure 3 and Supplementary Table 6**).

To further replicate our findings, we compared our SingleBrain microglia (MG) eQTLs with previously reported *cis*-eQTLs derived from primary microglia in three cohorts^8^ (Microglia Genomic Atlas study [MiGA]^5^, *Young* et al.^6^, and *Kosoy* et al.^7^). For a direct comparison, we reanalyzed these datasets using our eQTL pipeline, performing a meta-analysis—which we term “MiGA 3”. While the number of eGenes was comparable between SingleBrain MG (8,423) and MiGA 3 (20,084), a low degree of eQTL sharing between MiGA 3 and SingleBrain was observed (π_1_ = 0.34), suggesting that cellular and nuclear transcriptomic profiles might capture distinct eQTL signatures (**Supplementary Figure 5**).

### SingleBrain eQTLs mediate neuropsychiatric disease

To formally test whether there was evidence for sharing the same genetic effect between SingleBrain eQTLs and neurodegenerative and neuropsychiatric diseases, we calculated disease heritability using mediated expression score regression (MESC)^21^ and conducted colocalization and MR analyses, leveraging publicly available GWAS summary statistics for Alzheimer’s disease (AD)^22–25^, Parkinson’s disease (PD)^26^, schizophrenia (SCZ)^27^, multiple sclerosis (MS)^28^, bipolar disorder (BPD)^28^, and amyotrophic lateral sclerosis (ALS)^29^ (**Supplementary Figures 7-9, Supplementary Tables 7-10**).

We found that microglia eQTLs mediate the highest proportion of heritability for AD, while showing no enrichment for SCZ. PD exhibited non-zero heritability mediation across multiple cell types, suggesting broader cellular involvement. Except for AD, all brain disorders showed significant heritability mediation through neuronal eQTLs, with SCZ and BPD displaying the strongest enrichment (**Supplementary Figure 7**).

SCZ had the highest number of colocalizations, with 109 of 217 loci containing at least one colocalized gene based on a posterior probability (PP) of the causal variant being shared between eQTL and GWAS (PP4) > 0.8 (**Figure 3a**). MR provided additional evidence for causal association with disease outcomes for 58 of these loci and provided a direction of effect (**Figure 3a**). SCZ loci demonstrated the highest colocalization within neuronal cell types, particularly in excitatory (n = 46) and inhibitory neurons (n = 42) (**Figure 3b**). A small number of colocalizations in non-neuronal cell types such as the *CUL3* and *RERE* eGenes were found in astrocytes, as well as *YPEL4*, *SH3RF1*, and *SPIRE2* in microglia. Examples of other cell-type-specific colocalizations with SCZ are shown in **Figure 3c and Supplementary Figure 8**. In addition, we identified 26 colocalizations (out of 64 loci) with BPD, the majority of which were within neurons (**Figure 3b, Supplementary Figure 10, and Supplementary Table 11**).

**Figure 3.**
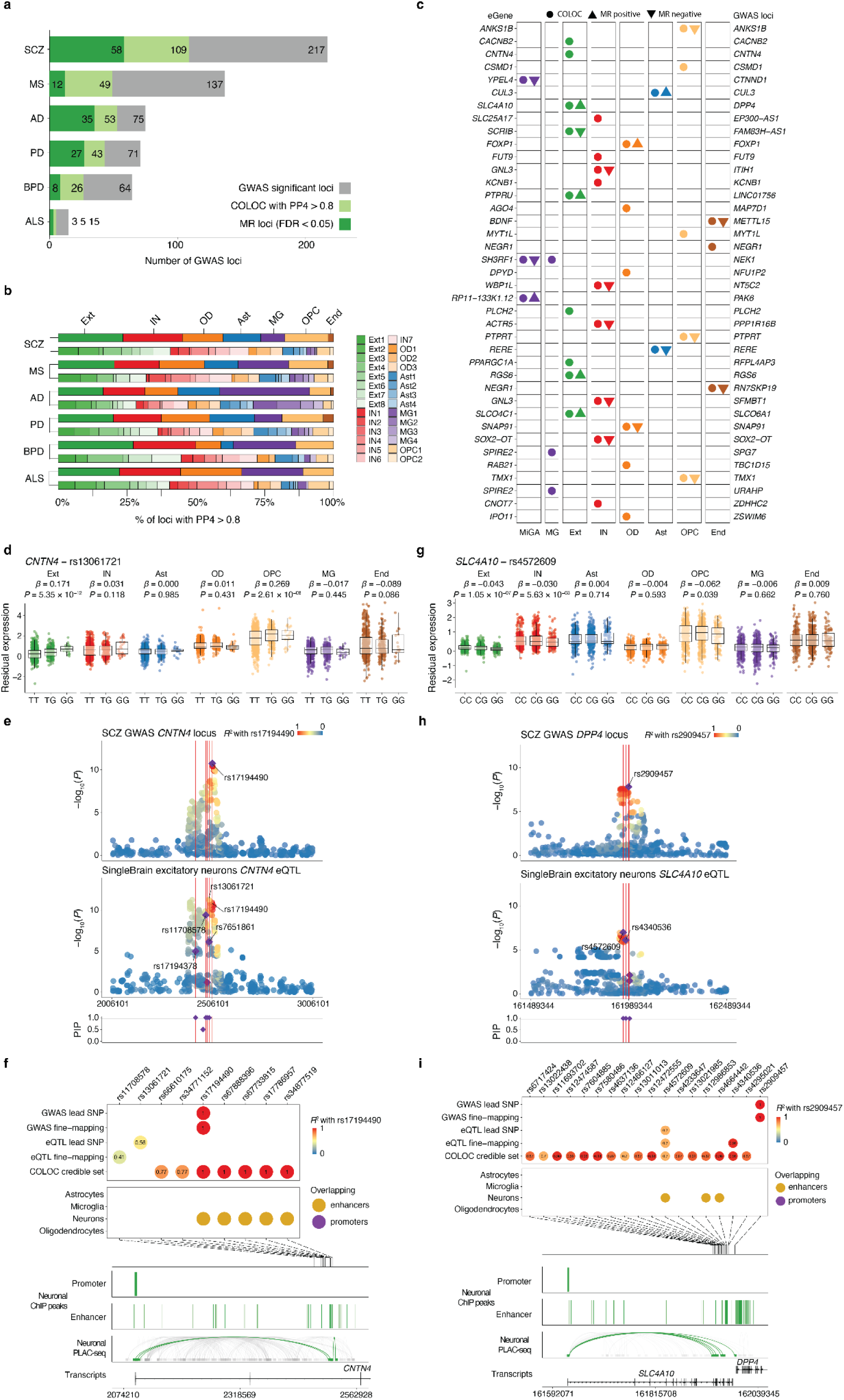
Colocalization of SingleBrain eQTLs with neurodegenerative and neuropsychiatric disease GWAS loci. **a,** Number of GWAS loci with at least one colocalization (probability of sharing one causal variant between eQTL and GWAS, PP4 > 0.8) and significant MR results (FDR < 0.05). The dark green indicates consistent loci between the COLOC and MR results. The light green indicates the COLOC (PP4 > 0.8) but a lack of evidence from MR. The gray indicates loci with no COLOC evidence (PP4 < 0.8). **b,** Percentage of GWAS loci that colocalize with cell-type- and subtype-specific eQTLs (PP4 > 0.8) across neurodegenerative and neuropsychiatric diseases. **c,** Cell-type-specific loci in schizophrenia (SCZ) GWAS that had a PP4 > 0.8 and significant MR association in SingleBrain and MiGA. The circle represents eQTL with PP4 > 0.8. The triangle denotes significant genes in MR results, with the orientation of the triangle corresponding to the direction of effect. **d-f,** Analysis of the *CNTN4* excitatory neurons eQTL with the SCZ GWAS *CNTN4* locus. **d,** *CNTN4* expression is associated with the rs13061721 genotype. **e,** Locus zoom and fine-mapping of *CNTN4* SCZ GWAS and *CNTN4* excitatory neurons eQTL. **f,** Fine-mapping of the *CNTN4* locus and combination with the GWAS lead SNP, fine-mapping SNP, eQTL lead SNP, fine-mapping SNP, and COLOC credible set SNPs of colocalization. **g-i,** Analysis of the *SLC4A10* excitatory neurons eQTL with the SCZ GWAS *DPP4* locus. **g,** *SLC4A10* expression is associated with the rs4572609 genotype. **h,** Locus zoom and fine-mapping of *DPP4* SCZ GWAS and *SLC4A10* excitatory neurons eQTL. **i,** Fine-mapping of the *DPP4* locus and combination with the GWAS lead and fine-mapping SNPs, eQTL lead and fine-mapping SNPs, and COLOC credible set SNPs of colocalization. **d,g,** The residual expression is PEER adjusted gene expression level. The nominal *P*-value and beta from the linear regression model in the brain cell type eQTL analysis are indicated above the box plots. The box plots show the median, the box spans from the first to the third quartiles, and the whiskers extend 1.5 times the interquartile range (IQR) from the box. **e,h,** Labels refer to lead SNPs and fine-mapping SNPs with *P* < 1 × 10^−4^. SNPs are colored by the LD with the lead GWAS SNP. Fine-mapping of GWAS and eQTL is present with a purple diamond under the locus zoom plot, and locus zoom only has a PIP > 0.95. **f,i,** SNPs are colored by the LD with the lead GWAS SNP. Cell-type enhancers and promoters were defined by *Nott* et al.^17^. Genomic plots (hg19) of the fine-mapped SNPs and epigenomic data from the microglia ChIP-seq and PLAC-seq junctions. SCZ, schizophrenia; MS, multiple sclerosis; AD, Alzheimer’s disease; PD, Parkinson’s disease; BPD, bipolar disorder; ALS, amyotrophic lateral sclerosis; COLOC, colocalization; MR, Mendelian randomization; Ext, excitatory neurons; IN, inhibihitory neurons; OD, oligodendrocytes; Ast, astrocytes; OPC, oligodendrocyte progenitor cells; MG, microglia; End, endothelial cells; MiGA, Microglia Genomic Atlas.

To further refine the regulatory mechanisms and pinpoint putative causal variants within each locus, we fine-mapped *cis*-eQTLs for each cell type **(Supplementary Table 12)**. Across all SCZ colocalizations, we were able to identify at least one 95% credible set for 76 loci (**Supplementary Figure 11**). Many of the SNPs in these fine-mapped credible sets reside within cell type-specific enhancers, emphasizing their potential regulatory roles. SCZ-associated eQTL for *CNTN4* in excitatory neurons shows that the protective allele rs13061721-G is linked to reduced *CNTN4* expression (**Figure 3d**). The fine-mapped SNPs, identified through COLOC credible set and GWAS lead SNPs, are within a neuron-specific enhancer. Proximity Ligation-Assisted ChIP-Seq (PLAC-Seq) data^17^, further revealed that these SNPs reside in a neuronal enhancer with extensive long-range interactions connecting to regions overlapping the promoter and gene body (**Figures 3e and 3f**). These findings suggest that risk variants enhance *CNTN4* expression through cell type-specific enhancers in excitatory neurons. One of the *CNTN4* COLOC credible set SNPs, rs6610175, has been validated in a recent massively parallel reporter assay (MPRA)^10^ (**Supplementary Figure 12 and Supplementary Table 13**).

Another example is the SCZ-associated *DDP4* locus, which colocalizes with the *SLC4A10* eQTL in neurons. The SCZ protective allele rs4572609-G is associated with lower *SLC4A10* expression(Figure 3g). The fine-mapped variant resides within neuron-specific enhancers and likely influences *SLC4A10* expression through the enhancers in excitatory neurons (Figures 3h and 3i). Interestingly, the *SLC4A10* eQTL exhibits subtype specificity, as colocalization is detected exclusively in the Ext2 (L4 IT3) subtype, with a PP4 of 0.84 (**Supplementary Figure 13**).

### SingleBrain eQTLs mediate neurodegenerative disease

We colocalized our sets of SingleBrain eQTLs with genome-wide significant loci in AD to identify mechanisms of risk mediation by effects on gene expression. Of the 75 genome-wide significant loci in the most recent AD GWAS^22^, 53 showed colocalization with SingleBrain eQTLs **(Figure 3a**). MR further confirmed 35 AD loci (FDR < 0.05) with significant association with gene expression, supporting causal links between genetic risk and gene regulation (**Figure 3b**). Consistent with our previous findings^5^, microglia eQTLs exhibited the highest number of colocalizations with AD GWAS loci (30 out of 75 GWAS loci) despite a lower overall number of eGenes (**Supplementary Figure 14**). Additionally, we observed colocalization with other cell types including 14 loci colocalized in astrocytes, 11 in oligodendrocytes, 15 in excitatory neurons, and 14 in inhibitory neurons, suggesting molecular mechanisms beyond microglia in AD pathogenesis (**Supplementary Figure 14**). *ABCA7 and TNIP1* colocalized exclusively in excitatory neurons, while *APP* and *EGFR* were specifically associated with astrocytes, highlighting distinct neuronal and glial contributions to AD. In microglia, we identified novel eQTL associations at *BLNK, RASA1, FERMT2, LILRB2, MS4A4E, LLGL1, NCK2* and *SORL1*, which were not detected in previous microglia or snRNASeq eQTL studies^5,11,12^ (**Figure 4a**). Finally, given the availability of multiple AD GWAS, we extended our colocalization analysis to include additional GWAS datasets^23–25^ (**Supplementary Figure 15)**.

**Figure 4.**
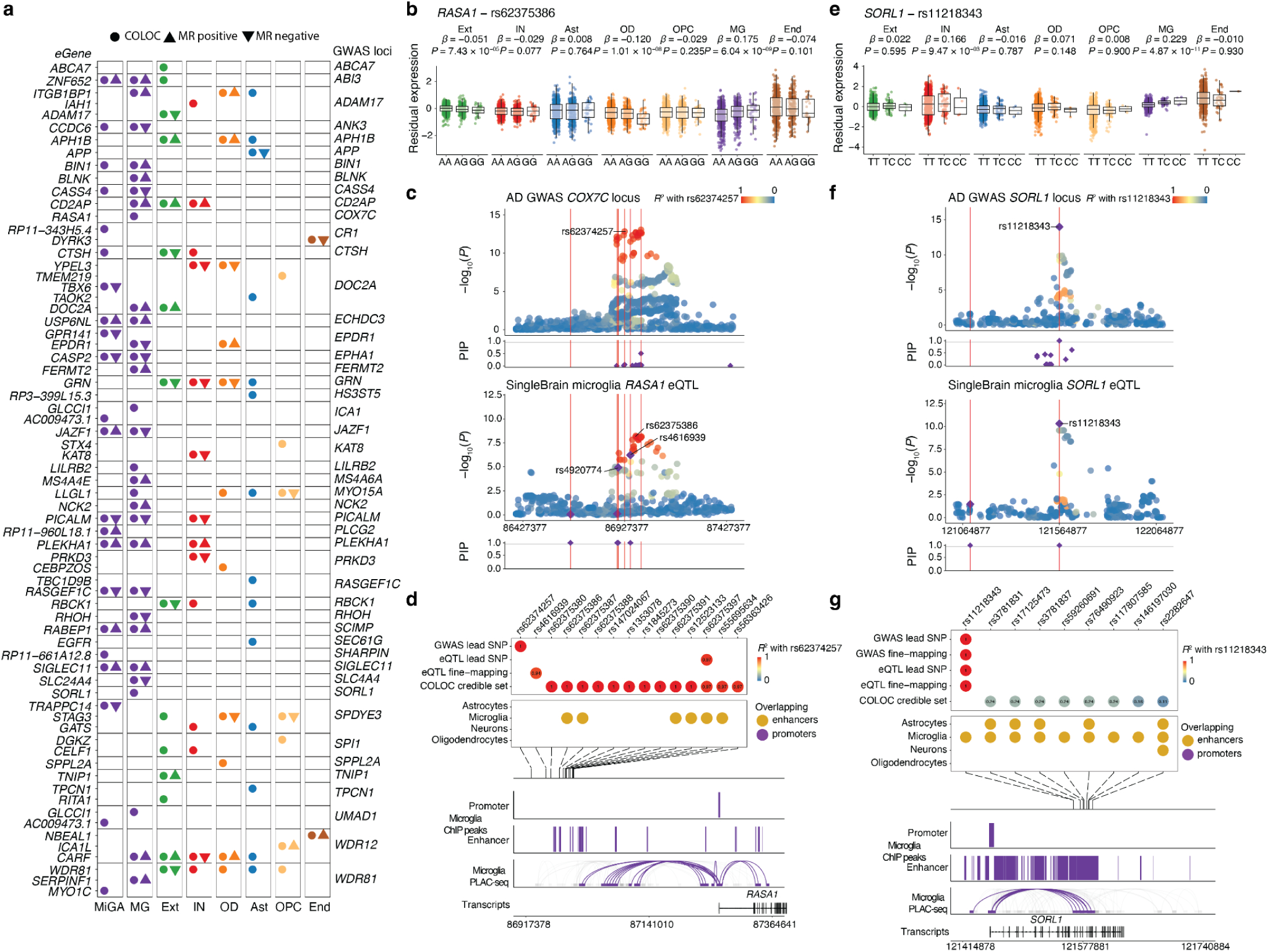
Microglia eQTLs are enriched in AD GWAS loci. **a,** All loci in Alzheimer’s disease (AD) GWAS that had a PP4 > 0.8 and significant MR association in SingleBrain and MiGA. The circle represents eQTL with PP4 > 0.8. The triangle denotes significant genes in MR results, with the orientation of the triangle corresponding to the direction of effect. **b-d,** Analysis of the *RASA1* microglia eQTL with the AD GWAS *COX7C* locus. **b,** *RASA1* expression is associated with the rs62375386 genotype. **c,** Locus zoom and fine-mapping of the *COX7C* AD GWAS and *RASA1* microglia eQTL. **d,** Fine-mapping of the *COX7C* locus and combination with the GWAS lead SNP, eQTL lead SNP, fine-mapping SNP, and COLOC credible set SNPs of colocalization. **e-g,** Analysis of the *SORL1* microglia eQTL with the AD GWAS *SORL1* locus. **e,** *SORL1* expression is associated with the rs11218343 genotype. **f,** Locus zoom and fine-mapping of the *SORL1* AD GWAS and *SORL1* microglia eQTL. **g,** Fine-mapping of the *SORL1* locus and combination with the GWAS lead SNP, fine-mapping SNP, eQTL lead SNP, fine-mapping SNP, and COLOC credible set SNPs of colocalization. **b,e,** The residual expression is PEER adjusted gene expression level. The nominal *P*-value and beta from the linear regression model in the brain cell type eQTL analysis are indicated above the box plots. The box plots show the median, the box spans from the first to the third quartiles, and the whiskers extend 1.5 times the interquartile range (IQR) from the box. **c,f,** Labels refer to lead SNPs and fine-mapping SNPs with *P* < 1 × 10^−4^. SNPs are colored by the LD with the lead GWAS SNP. Fine-mapping of GWAS and eQTL is present with a purple diamond under the locus zoom plot, and locus zoom only has a PIP > 0.95. **d,g,** SNPs are colored by the LD with the lead GWAS SNP. Cell-type enhancers and promoters were defined by *Nott* et al.^17^. Genomic plots (hg19) of the fine-mapped SNPs and epigenomic data from the microglia ChIP-seq and PLAC-seq junctions. COLOC, colocalization; MR, Mendelian randomization; MiGA; Microglia Genomic Atlas, MG, microglia; Ext, excitatory neurons; IN, inhibitory neurons; OD, oligodendrocytes; Ast, astrocytes; OPC, oligodendrocyte progenitor cells; End, endothelial cells.

To further pinpoint putative causal variants for each colocalized eQTL and GWAS locus, we conducted statistical fine-mapping of eQTLs. Fine-mapping of AD-associated eQTLs substantially refined the candidate causal variants, reducing the total from 1,173,104 to 1,420 SNPs with a PIP > 0.01 (**Supplementary Table 14**). Of these, 442 SNPs (31%) mapped within cell-type-specific enhancers, with the highest proportion, 128 (29%), residing in microglia-specific enhancers. These findings further highlight the central role of microglial regulatory elements in mediating AD genetic risk (**Supplementary Table 15**). For example, the *COX7C* AD risk locus colocalizes with microglial eQTLs for *RASA1*, which encodes the p120-RasGAP protein, with the alternative allele rs62375386-G (in LD with AD GWAS lead SNP, rs62374257) associated with increased expression (**Figure 4b**). The AD protective allele rs62374257-C (GWAS lead SNP), eQTL lead SNP and fine-mapped SNPs all overlap with microglia-specific enhancers, but not those of other cell types. PLAC-seq analysis revealed that this microglial enhancer region exhibits extensive long-range chromatin interactions with regions overlapping the *RASA1* promoter and gene body (**Figures 4c and 4d**), suggesting a potential regulatory mechanism linking genetic variation to *RASA1* expression in microglia.

Another notable example is an eQTL at the AD-associated *SORL1* locus, where the AD-risk allele rs11218343-C increases *SORL1* expression in microglia (**Figure 4e**). GWAS, eQTL, and COLOC credible set SNPs all localize within a microglia-specific enhancer. PLAC-seq data reveal extensive long-range interactions between this enhancer and the *SORL1* promoter (**Figures 4g and 4g**). This variant regulates *SORL1* expression via a microglia enhancer, with strong colocalization (PP4 = 0.95) in MG1 (a subtype enriched with surveillance genes^20^) (**Supplementary Figure 16**). Both *RASA1* and *SORL1* were likely undetected in our previous MiGA study due to the greater statistical power provided by the expanded SingleBrain microglia nuclei dataset (**Supplementary Figure 17**).

Of the 76 PD GWAS loci, 43 colocalized with SingleBrain eQTLs (PP4 > 0.8), and MR identified significant associations for 27 of these loci (FDR < 0.05) (**Figure 3b**). The majority of the PD GWAS loci (30) colocalized with neuronal eQTLs including many not discovered in other eQTL studies including *ASXL3, CLCN3* and *FCF1* in excitatory neurons and *CLCN3* in inhibitory neurons. We also identified many eQTLs that mediate PD risk in non-neuronal cells, including 14 eGenes colocalized in oligodendrocytes, 13 in astrocytes, 12 in OPCs, and 8 in microglia (**Figure 5a and Supplementary Figure 14**). Astrocyte-specific colocalization included *MED13*, *CD38*, *SETDB2*, *KCNIP3* and *STK39*. In oligodendrocytes, *TOX3* was identified, while *ITGA8*, *MIPOL1*, *PPIP5K2*, and *SYT17* were specific to OPCs (**Figure 5a**).

**Figure 5.**
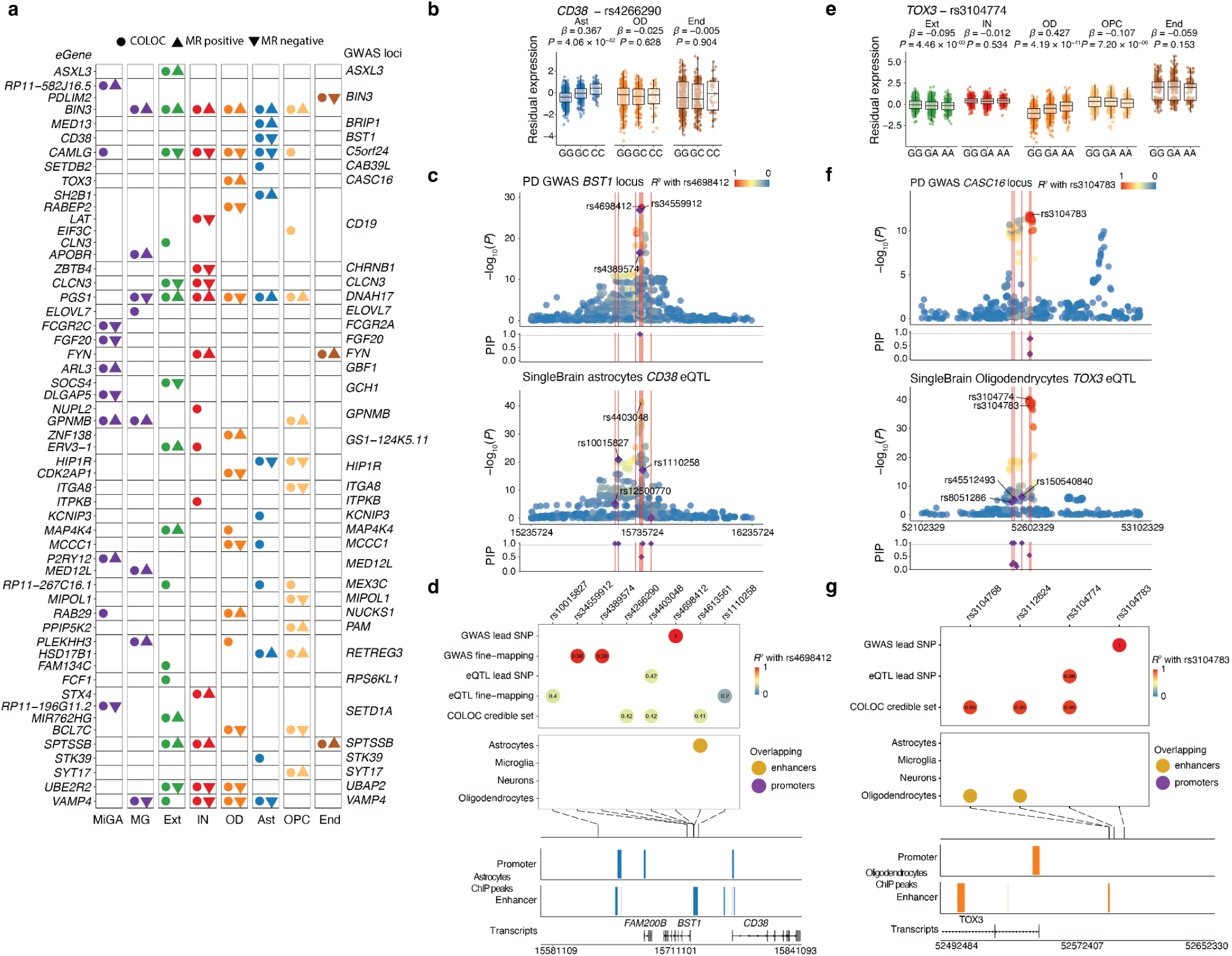
Brain cell-type-specific eQTLs drive genetic risk in PD. **a,** All loci in Parkinson’s disease (PD) GWAS that had a PP4 > 0.8 and significant MR association in SingleBrain and MiGA. The circle represents eQTL with PP4 > 0.8. The triangle denotes significant genes in MR results, with the orientation of the triangle corresponding to the direction of effect. **b-d,** Analysis of the *CD38* astrocytes eQTL within the *BST1* locus. **b,** *CD38* expression is associated with the rs4266290 genotype. **c,** Locus zoom and fine-mapping of the *BST1* PD GWAS and *CD38* astrocyte eQTL. **d,** Fine-mapping of the *BST1* locus and combination with the GWAS lead SNP, fine-mapping SNPs, eQTL lead SNP, fine-mapping SNPs, and COLOC credible set SNPs overlaid with astrocyte epigenomic data. **e-g,** Analysis of the *TOX3* oligodendrocytes eQTL with the PD GWAS *CASC6* locus. **e** *TOX3* expression is associated with the rs3104774 genotype. **f,** Locus zoom and fine-mapping of the *CASC6* PD GWAS and *TOX3* oligodendrocytes eQTL. **g,** Fine-mapping of the *CASC6* locus and combination with the GWAS lead SNP, QTL lead SNP, and COLOC credible set SNPs overlaid with oligodendrocyte epigenomic data. **b,e,** Expression level residualized by PEER factors. The nominal *P*-value and beta from the linear regression model in the brain cell type eQTL analysis are indicated above the box plots. The box plots show the median, the box spans from the first to the third quartiles, and the whiskers extend 1.5 times the interquartile range (IQR) from the box. **c,f,** Labels refer to lead SNPs and fine-mapping SNPs with *P* < 1 × 10^−4^. SNPs are colored by the LD with the lead GWAS SNP. Fine-mapping of GWAS and eQTL is present with a purple diamond under the locus zoom plot, and locus zoom only has a PIP > 0.95. **d,g,** SNPs are colored by the LD with the lead GWAS SNP. Cell-type enhancers and promoters were defined by *Nott* et al.^17^. COLOC, colocalization; MR, Mendelian randomization; MiGA; Microglia Genomic Atlas, MG, microglia; Ext, excitatory neurons; IN, inhibitory neurons; OD, oligodendrocytes; Ast, astrocytes; OPC, oligodendrocyte progenitor cells; End, endothelial cells.

An example of a non-neuronal eQTL is observed at the *BST1* locus, where the PD protective allele rs4266290-C increases *CD38* expression specifically in astrocytes **(Figure 5b)**. The lead PD GWAS SNP, rs4698412 (in LD with the lead eQTL, R² < 0.42), is in an astrocyte-specific enhancer **(Figure 5c and 5d).** Another example is an eQTL at the PD-associated *CASC6* locus, where the PD-risk allele rs3104774-A increases *TOX3* expression in oligodendrocytes (**Figure 5e**). The COLOC credible set variants fall within an oligodendrocytes-specific enhancer. These variants influence *TOX3* expression through the enhancer in the oligodendrocytes cell type (**Figures 5f and 5g**).

### Microglia subtypes-specific eQTLs

Microglia exist in a wide range of dynamic states, reflecting their diverse functional roles in the CNS^30^. Building on previous studies^15,20^, we identified distinct microglial subtypes and performed subtype-specific *cis*-eQTL analyses^20^. Although up to 16 microglial subpopulations have been previously reported^31,32^, we focused our subpopulation eQTL analysis on four microglial populations due to the limited power and low number of nuclei per donor for rarer microglial populations. The four major subtypes—MG1, MG2, MG3, and MG4—correspond to the clusters identified by Green *et al.*^20^ For microglia subtypes, we projected our clusters as follows: MG1 (Mic 2-5), containing surveilling genes (*CXCR1*⁺, *P2RY12*⁺); MG2 (Mic6-8), containing reactive genes (*TMEM163*⁺, *IL4R*⁺); MG3 (Mic9-10), enriched for redox-related genes; and MG4 (Mic12-13), characterized by lipid-processing genes (*APOE*⁺, *ADAM10*⁺, *GPNMB*⁺). For these subtypes, we were able to identify and colocalize eQTLs with AD and PD GWAS loci, providing insights into the genetic control of gene expression within subtypes (**Figure 6a and Supplementary Figure 16**). While many eQTLs detected in pseudobulk microglia were shared across multiple states, we identified several subtype-specific eQTLs (**Supplementary Figure 14**). For example, at the AD loci *SORL1* and *PARP1*, we identified colocalization with eQTLs specifically in the MG1 subtype, but not in other microglial subtypes (**Figure 6a and 6b**). For PD, we identified a greater number of subtype-specific eQTLs in microglia. We found several PD-associated eQTLs unique to specific microglial subtypes, including *TMEM120B* in MG1, *FGF20* in MG3, *GPR65* in MG4, and *SH3RF1* in both MG3 and MG4 (**Figure 6c and Supplementary Figure 18**).

**Figure 6.**
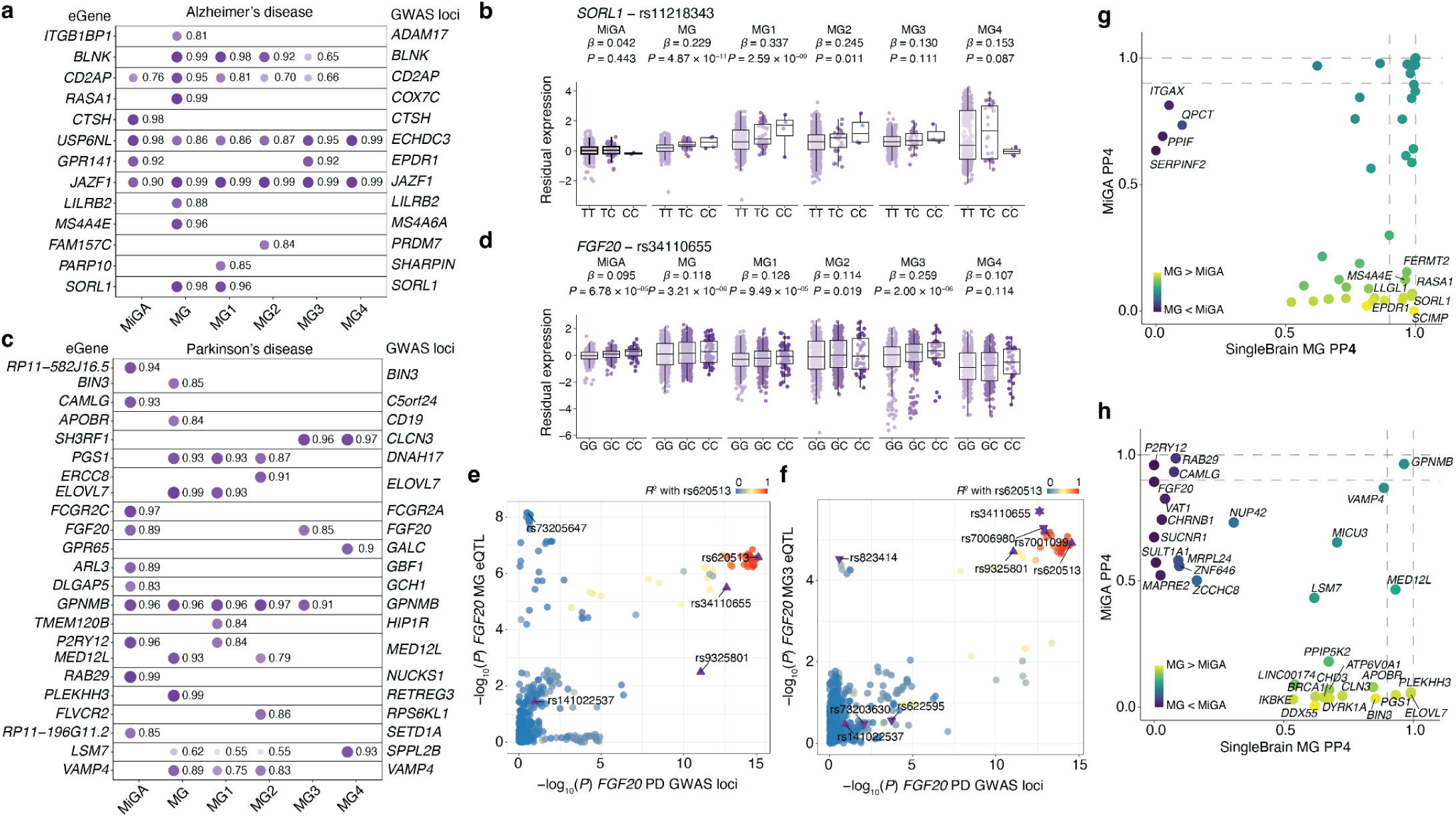
Disease-associated eQTLs in microglia subtypes. **a,** Subtype-specific AD GWAS locus had a PP4 > 0.8 colocalization at SingleBrain microglia subtypes and MiGA. Shape opacity and size were scaled to the magnitude of PP4. Numbers next to the circles are PP4 values. **b,** *SORL1* expression) is associated with the rs11218343. **c,** All PD GWAS locus that had a PP4 > 0.8 colocalization in SingleBrain microglia subtypes and MiGA. Value is PP4. Shape opacity and size are scaled to PP4. **d,** *FGF20* expression is associated with rs34110655. **e**, The *FGF20* PD GWAS loci with *FGF20* MG (pseudobulk) eQTL. For **b,** and **d,** residual expression is PEER adjusted gene expression level. The nominal *P*-value and beta from the linear regression model in the brain cell type eQTL analysis are indicated above the box plots. The box plots show the median, the box spans from the first to the third quartiles, and the whiskers extend 1.5 times the interquartile range (IQR) from the box. **f**, The *FGF20* PD GWAS locus with *FGF20* MG3 eQTL. SNPs are colored by the LD with the lead GWAS SNP. The triangle indicates GWAS fine-mapping SNPs and the inverted triangle indicates the eQTL fine-mapping SNPs (PIP > 0.95). **g,** Comparison of the maximum SingleBrain microglia and MiGA PP4 for each AD locus across both references. Labels refer to selected loci. **h,** Comparing the maximum SingleBrain microglia and MiGA PP4 for each PD locus across both references. MiGA, Microglia Genomic Atlas; MG, microglia.

An example is the *FGF20* eQTL (PP4 = 0.85), which is significant in MG3 but not in other subtypes (**Figure 6d-6f, Supplementary Figures 18 and 19**), highlighting the specificity of eQTL effects within distinct microglial populations. GWAS and eQTL fine-mapped SNPs all overlap with microglia-specific enhancers. Using the PLAC-seq, this microglial enhancer region shows long-range chromatin interactions with regions overlapping the *FGF20* promoter and gene body (**Supplementary Figure 20**), suggesting a potential regulatory mechanism linking genetic variation to *FGF20* expression in MG3.

Lastly, we compared microglia eQTLs identified in SingleBrain with those from purified microglia cells in MiGA 3. Although we observed overlapping disease-associated colocalizations for AD and PD in both datasets, several eQTLs were uniquely detected in either SingleBrain or MiGA (**Figure 6g, 6h and Supplementary Table 16**). This highlights distinct regulatory landscapes between whole cells and isolated nuclei, emphasizing the importance of cellular context in understanding gene regulation.

## Discussion

We present SingleBrain, a meta-analysis of snRNA-seq eQTLs in the human brain, integrating data from four major cohorts comprising 5.8 million nuclei from 983 donors. This significantly expands the landscape of brain eQTLs across seven major cell types and 28 subtypes. Compared to single-cohort studies, SingleBrain identifies a 3–10-fold increase in *cis*-eQTLs, enabled by its larger sample size and high-resolution data. This approach enhances eQTL discovery in abundant cell types, such as excitatory and inhibitory neurons, and enables detection in rare populations, including microglia and endothelial cells, previously limited by statistical power. Finally, we conducted statistical fine-mapping of SingleBrain eQTLs and neurodegenerative and neuropsychiatric disease GWAS to prioritize putative causal variants at each locus and integrated analyses to map cell type-specific enhancer-promoter regulatory interactions.

Our analysis reveals cell type-specific genetic regulatory mechanisms underlying major neuropsychiatric and neurodegenerative disorder, identifying 143 loci that significantly colocalize with singleBrain eQTLs based on COLOC (PP4 > 0.8) and MR analyses (FDR < 0.05). As expected, we observed the most colocalization signals between microglia eQTLs and AD risk loci, while neuronal eQTLs showed colocalization with risk loci for SCZ, BPD, and PD. We also identified several SCZ, BPD, and PD-associated loci that colocalize with eQTLs in oligodendrocytes, astrocytes, and microglia. Additionally, the extensive number of nuclei analyzed provided sufficient power to resolve cell subtype-specific eQTLs, revealing novel regulatory effects within distinct neuronal subtypes and specific cortical layers that were not previously detected.

Our meta-analysis of 260,441 microglia nuclei enabled high-resolution, subtype-specific eQTL analyses. The subtype specific eQTL analysis identified many novel eQTLs including *SORL1* which was detected exclusively in MG1, reinforcing the idea that AD genetic risk may be mediated through specific microglial subpopulations. *FGF20* eQTLs at the PD GWAS loci were detected only in MG3, highlighting distinct genetic influences on microglial subtypes and their roles in neurodegeneration. These findings suggest that genetic variants exert context-specific regulatory effects based on microglial activation, inflammation, or disease progression, emphasizing the importance of considering microglial state dynamics in eQTL interpretation.

We also discovered a subset of microglial eQTLs detectable only in purified microglial cells (MiGA 3) but not in SingleBrain nuclei MG, and vice versa. These findings highlight the complementary nature of these approaches and the potential influence of differences in data types, methodologies, and sample characteristics. Several potential factors could explain this discrepancy. The lower abundance of certain genes in nuclear RNA compared to whole-cell RNA can hinder their detection in snRNA-seq data, particularly for genes with low nuclear expression or those depleted in nuclei preparation^33^. Additionally, technical limitations, such as restricted read depth or an insufficient number of cells per sample in SingleBrain microglia datasets, may further constrain eQTL identification. These challenges underscore the need for integrative strategies that incorporate both nuclear and whole-cell datasets for a more comprehensive mapping of eQTLs across microglial states.

Despite its strengths, this study has several limitations. First, snRNA-seq primarily captures nuclear RNA which may not fully represent the cytoplasmic transcriptome. Furthermore, while 10x 3’ libraries are designed to best capture the 3’ ends of genes, cDNAs are primed throughout genes, particularly within introns. Gene quantification in nuclei typically combines exonic and intronic read counts over the entire body of the gene to maximize gene detection, at the expense of potentially misassigning reads to overlapping genes. Combining spliced and unspliced reads for eQTLs mapping would also have the potential confounding factor of picking up pre-mRNAs. Certain genes with low nuclear expression or unique cytoplasmic regulation may be underrepresented, limiting eQTL detection for these transcripts. Second, rare cell types and microglial subtypes had limited nuclei per donor, reducing statistical power to detect eQTLs in these groups. Future studies with larger sample sizes or enriched datasets for rare populations could address this limitation. Lastly, our analysis was restricted to donors of European ancestry, which limits the generalizability of our findings to other populations. Expanding this research to diverse ancestral backgrounds will be critical to ensure inclusivity and broader applicability.

SingleBrain represents a significant step forward in understanding the genetic regulation of brain cell transcriptomes and its implications for neurological and neuropsychiatric disorders. By providing a high-resolution map of eQTLs across diverse brain cell types and states, this resource advances our understanding of the molecular mechanisms underlying complex brain diseases. Importantly, we prioritize putative genes and genetic variants for further mechanistic investigations, laying the groundwork for future functional studies. Future efforts to integrate this dataset with other omics layers, increase sample diversity, and perform experimental validations will further enhance the utility and impact of this resource.

## Methods

### Single Nucleus RNA-seq cohorts

We collected published genotypes and raw count matrices of single-nucleus RNA-seq samples from *Fujita* et al.^11^, *Mathys* et al.^14^, *Gabitto* et al.^15^ from Synapse.org using the Python 3.8.5 package synapseclient v.4.5.1^34^. and the Roche datasets have been deposited at the European Genome-phenome Archive, which the European Bioinformatics Institute and the Centre for Genomic Regulation host^12^. For all datasets^11,12,14,15^, annotated cells and samples as poor-quality were excluded. The four snRNA-seq datasets included in SingleBrain comprised 1,047 samples from 983 donors, including 757 unique donors. (**Supplementary Table 1**). *Fujita* et al. and *Mathys* et al. cohorts shared 226 overlapping donors.

### Genotype Data Quality Control

#### Whole-genome sequencing

To perform genotype quality control (QC), we used the Python library Hail v.0.2.113^35^. Multi-allelic variants were split into biallelic variants before QC. Variants with low Variant Quality Score Recalibration quality or located within known low-complexity regions were filtered out. Sample-level QC removed samples with low mean depth, low call rate, or discrepancies between genotype and sex. Non-European samples were excluded using Somalier^36^ v.0.2.12. In the European population samples, outliers with a variant count outside 4 or 8 standard deviations (SDs) from the mean were removed. After sample-level QC, we filtered out variants that had low variant call rates, failed the Hardy-Weinberg equilibrium test, or had low variant quality scores for each single-nucleotide polymorphism (SNP) and insertion and deletion (INDEL). Detailed information on genetic QC is provided in **Supplementary Methods**.

#### Chip array genotypes

We performed QC of SNP data. SNPs were kept using the following criteria: (i) missing rate of < 2%, (ii) minor allele frequency (MAF) of >1%, or (iii) Hardy–Weinberg equilibrium (HWE) with a *P* value of >10^−6^. Based on the genotype data, participants were included following criteria: (i) missing rate of <1%, (ii) sex match, (iii) heterozygosity excess (± 3 standard deviations from the mean), or (iv) European ancestry of samples was estimated with Somalier^36^ v.0.2.12.

Imputation was conducted using the Minimac4 software with all available reference haplotypes from the Trans-Omics for Precision Medicine (TOPMed) at the Michigan Imputation Server. Consequently, we performed post-imputation QC with (i) an MAF of >1% or (ii) a high imputation quality (*r*^2^ > 0.8 for imputed SNPs). Genome annotations were generated using the GRCh38 assembly.

### Single-nuclei RNA-seq analysis

To process single-cell RNA sequencing, we used SCANPY^37^ (v1.9.8). First, we excluded outlier cells (3 x the IQR) in terms of the number of genes, total counts, and percentage of mitochondrial genes. Second, we removed doublets detected using Scrublet^38^ (v.0.2.3). We removed single-cell platform and dataset-specific batch effects using Harmony^39^, using individual and batches with normalized gene expression. The major cell types were then annotated based on annotations from original studies^11,12,14,15^. Based on previously published single-cell RNA-sequencing data, subtypes projected annotations with highly variable 5,000 genes by each major cell type. The neurons, oligodendrocytes, and OPCs defined by *Gabitto* et al.^15^ were mapped onto this study. We leveraged *Green* et al.^20^ cell and subtype annotations to project and refine the classification of astrocytes and microglia, aligning them with our identified clusters. All major cell type and subtype abbreviations are defined in **Supplementary Table 6**.

### Pseudobulk expression quantification

Pseudo-bulk gene expression matrices were aggregated in all counts for each gene in each individual by each cell type. Individuals contributing fewer than ten cells in a cell type were filtered out. Genes with low expression, defined as those with more than 10% of cells showing zero expression, were excluded from the analysis. Rare cell types were excluded from subsequent analysis if fewer than 20 individuals were available for expression quantification for each dataset. Using edgeR (v.3.40.2) with its default parameters, the trimmed mean of M-values method was used to normalize the pseudobulk counts. To compute the log_2_ of counts per million mapped reads (CPM), we used the limma (v.3.54.2) voom function. Less than 2.0 log_2_ CPM was filtered out as lowly expressed genes. Pseudobulk expression of cell subtypes was quantified using the same method.

### Mapping of *cis*-eQTL and meta-analysis

To map QTLs and perform a meta-analysis, we used mmQTL^16^ v.26a. First, with a MAF cutoff of 0.01, for each SNP–gene pair within a 1-Mb upstream of start of the feature and 1 Mb downstream of end of feature window, mmQTL applies a linear mixed model with PEER residualized expression for QTL mapping. We used PEER^40^ factors to correct the batch effect and other covariates. For the major brain cell types, the number of PEER factors was chosen using GTEx as a guide^41^, with sample sizes of <150, 150-250, and >250 being assigned 15, 30, 35 PEER factors, respectively. Pseudobulk expression of subtypes was quantified using the same method, and 5 PEER factors were used.

To account for population stratification, the genotype relatedness matrix constructed using GCTA^42^ v.1.94.1 after converting to Plink^43^ (v.2.3) format was included to estimate QTL effect size in the linear mixed model. After QTL mapping for each dataset, the resultant effect sizes^44^ were combined in a random-effects meta-analysis using mmQTL^16^ v.26a. *P*-values are first FDR adjusted for the number of SNPs tested with each phenotype, then corrected for the number of phenotypes tested using Storey’s q-value^45^.

### GWAS summary stats

We obtained publicly available GWAS summary statistics for AD (*Marioni* et al.^25^, *Jansen* et al.^24^, *Kunkle* et al.^23^, *Bellenguez* et al.^22^), ALS (*van Rheenen* et al.^29^), BPD (*Mullins* et al.^46^, MS (*IMSGC* et al.^28^), and SCZ (*Trubetskoy* et al.^27^) and PD (*Nalls* et al.^26^). For each GWAS, we downloaded the full summary statistics and a list of genome-wide significant loci, as defined separately by each study. Missing fields in the nominal statistics were dealt with as follows: the standard error was calculated from the effect size and P value; minor allele frequency (MAF) was taken from European ancestry samples from gnomAD v4^47^; SNP coordinates or rsIDs were matched using Ensembl release 99.

### Colocalization

We used the COLOC package (v.3.2-1). We extracted a significant genome-wide locus within 1 Mb on either side of the lead SNP (2 Mb-wide region total) in the GWAS. In each QTL dataset, we filtered all SNPs by each gene matched with a significant genome-wide locus within 100 kb to test for colocalization. Missing minor allele frequency was used by reference values from the gnomAD EUR superpopulations^47^. For each locus, we tested co-localization between the GWAS and eQTL signals for genes with at least ten SNPs. Matching sets of SNPs colocalized by comparing the *P*-value distributions. We restricted the GWAS and QTL lead SNP pairs to those with distances ≤500 kb or in moderate LD (*r*^2^ > 0.1) using the 1000 Genomes (Phase 3) CEU population calculated in the LDLinkR package^48^ (v.1.1.2). The COLOC credible set are SNPs with a cumulative PP4 greater than 95%.

### Summary-data-based Mendelian Randomization

To perform the Mendelian randomization, we used the TwoSampleMR^49^ R package (v.0.6.8) with cell-type eQTL effect sizes by gene as an exposure and GWAS for a given neurological disease as an outcome. Genes are selected with a colocalization PP4 > 0.8. As an instrumental variable, the eQTL SNPs were filtered by FDR below 0.05. Then, we conducted the clumping of the variants using LD from 1000 Genome Projects European panels^50^ (*r*^2^ >0.01) within 1Mbp. Missing minor allele frequency was used by reference values from the gnomAD EUR superpopulations^47^. We mainly used the inverse variance weighted (IVW)^51^ method to analyze the dataset after filtering the FDR below 0.05 calculated by Benjamini-Hochberg in each GWAS. Other methods including Weighted median^52^ and MR Egger^53^ are provided in **Supplementary Table 10**.

### eQTL fine-mapping

We performed statistical and functional fine-mapping of each eQTL with a suggestive colocalization PP4 > 0.5 by each cell type. The full 1Mb window was fine-mapped in each locus to better take into account more widespread LD architectures. SNPs with minor allele frequency (MAF) < 1% were removed as we were primarily focused on identifying common risk factors. We fine-mapped using POLYFUN + FINEMAP and POLYFUN + SUSIE^54^, with LD from UKBiobank. LD correlation matrices (in units of *r*) were acquired for each eQTL loci from the UK Biobank (UKB) reference panel, pre-calculated by *Weissbrod* et al.^55^. We set the maximum number of causal SNPs per eQTL to 5. Each tool produces a 95% credible set of SNPs, which can be understood as SNPs with a posterior probability > 95% of being causal for the given phenotype.

### Epigenomic data integration

To prioritize genes and genetic variants, we used the processed cell-type specific promoter and enhancer data (*Nott* et al.^17^). Briefly, fluorescent activated nuclear sorting (FANS) was performed on postmortem human brains to isolate PU.1+ microglia, NEUN+ neurons, OLIG2+ oligodendrocytes, and NEUN-/LHX2+ astrocyte nuclei. Chromatin immunoprecipitation sequencing (ChIP-seq) was performed for the histone modifications H3K27ac and H3K4me3, identifying activated chromatin regions and promoters, respectively. In addition, proximity ligation-assisted ChIP-seq (PLAC-seq)^56^ was performed to identify long-range connections between H3K4me3-positive promoter regions and other genomic regions. To map between our SNPs set in each GWAS locus, ChIP-seq and PLAC-seq were coordinated by each cell type using GenomicRanges R packages^57^.

## Data availability

Genotypes from Fujita et al. and Mathys et al. are available at https://doi.org/10.7303/syn10901595. snRNA-seq data from *Fujita* et al. are available at https://www.synapse.org/Synapse:syn31512863. snRNA-seq data from *Mathys* et al. are available at https://www.synapse.org/Synapse:syn52293417. snRNA-seq and genotype data from *Gabitto* et al. are available at https://www.synapse.org/Synapse:syn26223298. Single-nuclei RNA-seq data and genotype data for the Roche cohort, which is part of *Bryois* et al., have been deposited at the European Genome-phenome Archive, which is hosted by the European Bioinformatics Institute and the Centre for Genomic Regulation, under accession number EGAS00001006345.

MPRA schizophrenia dataset was downloaded at https://github.com/thewonlab/schizophrenia-MPRA. Epigenomic data from purified human microglia, neurons, astrocytes, and oligodendrocytes^17^ were downloaded at https://github.com/nottalexi/brain-cell-type-peak-files.

SingleBrain eQTL browser is accessible at https://singlebrain.nygenome.org/eqtl/. All QTL summary statistics are accessible at https://zenodo.org/records/14908182.

## Code availability

QTL preparation and meta-analysis pipeline: https://github.com/RajLabMSSM/mmQTL-pipeline. Downstream-QTL analysis pipeline: https://github.com/RajLabMSSM/downstream-QTL.

All code used to produce analysis for figures: https://github.com/RajLabMSSM/SingleBrain.

## Acknowledgments

We thank the patients and families who donated material for these studies. T.R. and J.B. received funding from Calico Life Sciences LLC, and T.R., B.J., W.H.C., T.N., J.H. was supported by National Institutes of Health (NIH) grants, including NIA U01-AG058635, NIA R21-AG063130, NIA R01-AG054005, NIA U01-AG068880, NIA RF1-AG065926, NIA R56-AG055824, NIA P30-AG066514, NINDS U54-NS123743, and NINDS R01-NS116006. H.-H.W. was supported by the Ministry of Health & Welfare, Ministry of Science and ICT, Republic of Korea (RS-2022-KH125557), the National Research Foundation of Korea (NRF) (RS-2023-00223277), and by the Future Medicine 2030 Project of the Samsung Medical Center (#SMX1250081).

This work was supported in part through the computational resources and staff expertise provided by Scientific Computing at the Icahn School of Medicine at Mount Sinai and supported by the Clinical and Translational Science Awards (CTSA) grant UL1TR004419 from the National Center for Advancing Translational Sciences. Research reported in this paper was supported by the Office of Research Infrastructure of the National Institutes of Health under award number S10OD026880 and S10OD030463.

## Author contributions

T.R. conceived and designed the study. B.J. analyzed the data and performed the statistical analyses with assistance from K.B.P., W.H.C., A.R., S.-H.J., T.N., B.K., M.S.K., M.C., M.-S.P., M.R., and J.H. and with supervision by H.-H.W. and T.R.. A.T. constructed the website. B.J., H.-H.W., and T.R. interpreted the results. B.J. and T.R. wrote the manuscript, and J.H., D.A.K., and H.-H.W revised the manuscript; all authors have read and approved the final manuscript. T.R. and H.-H.W. supervised the project.

## Ethics declarations Competing interests

The authors declare no conflicts of interest for this study. T.R. served as a scientific advisor for Merck and serves as a consultant for Curie.Bio.

## References

1. Ng, B. et al. An xQTL map integrates the genetic architecture of the human brain’s transcriptome and epigenome. Nat. Neurosci. 20, 1418–1426 (2017).

2. de Klein, N. et al. Brain expression quantitative trait locus and network analyses reveal downstream effects and putative drivers for brain-related diseases. Nat. Genet. 55, 377–388 (2023).

3. GTEx Consortium. The GTEx Consortium atlas of genetic regulatory effects across human tissues. Science 369, 1318–1330 (2020).

4. Fromer, M. et al. Gene expression elucidates functional impact of polygenic risk for schizophrenia. Nat. Neurosci. 19, 1442–1453 (2016).

5. Lopes, K. de P., et al. Genetic analysis of the human microglial transcriptome across brain regions, aging and disease pathologies. Nat. Genet. 54, 4–17 (2022).

6. Young, A. M. H. et al. A map of transcriptional heterogeneity and regulatory variation in human microglia. Nat. Genet. 53, 861–868 (2021).

7. Kosoy, R. et al. Genetics of the human microglia regulome refines Alzheimer’s disease risk loci. Nat. Genet. 54, 1145–1154 (2022).

8. Humphrey, J. et al. Long-read RNA-seq atlas of novel microglia isoforms elucidates disease-associated genetic regulation of splicing. medRxiv (2023) doi:10.1101/2023.12.01.23299073.

9. Jerber, J. et al. Population-scale single-cell RNA-seq profiling across dopaminergic neuron differentiation. Nat. Genet. 53, 304–312 (2021).

10. McAfee, J. C. et al. Systematic investigation of allelic regulatory activity of schizophrenia-associated common variants. Cell Genom. 3, 100404 (2023).

11. Fujita, M. et al. Cell subtype-specific effects of genetic variation in the Alzheimer’s disease brain. Nat. Genet. 56, 605–614 (2024).

12. Bryois, J. et al. Cell-type-specific cis-eQTLs in eight human brain cell types identify novel risk genes for psychiatric and neurological disorders. Nat. Neurosci. 25, 1104–1112 (2022).

13. Emani, P. S. et al. Single-cell genomics and regulatory networks for 388 human brains. Science 384, eadi5199 (2024).

14. Mathys, H. et al. Single-cell atlas reveals correlates of high cognitive function, dementia, and resilience to Alzheimer’s disease pathology. Cell 186, 4365–4385.e27 (2023).

15. Gabitto, M. I. et al. Integrated multimodal cell atlas of Alzheimer’s disease. Nat. Neurosci. 27, 2366–2383 (2024).

16. Zeng, B. et al. Multi-ancestry eQTL meta-analysis of human brain identifies candidate causal variants for brain-related traits. Nat. Genet. 54, 161–169 (2022).

17. Nott, A. et al. Brain cell type-specific enhancer-promoter interactome maps and disease-risk association. Science 366, 1134–1139 (2019).

18. Dimas, A. S. et al. Common regulatory variation impacts gene expression in a cell type-dependent manner. Science 325, 1246–1250 (2009).

19. Gamazon, E. R. et al. Using an atlas of gene regulation across 44 human tissues to inform complex disease- and trait-associated variation. Nat. Genet. 50, 956–967 (2018).

20. Green, G. S. et al. Cellular communities reveal trajectories of brain ageing and Alzheimer’s disease. Nature 633, 634–645 (2024).

21. Yao, D. W., O’Connor, L. J., Price, A. L. & Gusev, A. Quantifying genetic effects on disease mediated by assayed gene expression levels. Nat. Genet. 52, 626–633 (2020).

22. Bellenguez, C. et al. New insights into the genetic etiology of Alzheimer’s disease and related dementias. Nat. Genet. 54, 412–436 (2022).

23. Kunkle, B. W. et al. Genetic meta-analysis of diagnosed Alzheimer’s disease identifies new risk loci and implicates Aβ, tau, immunity and lipid processing. Nat. Genet. 51, 414–430 (2019).

24. Jansen, I. E. et al. Genome-wide meta-analysis identifies new loci and functional pathways influencing Alzheimer’s disease risk. Nat. Genet. 51, 404–413 (2019).

25. Marioni, R. E. et al. GWAS on family history of Alzheimer’s disease. Transl. Psychiatry 8, 99 (2018).

26. Nalls, M. A. et al. Identification of novel risk loci, causal insights, and heritable risk for Parkinson’s disease: a meta-analysis of genome-wide association studies. Lancet Neurol. 18, 1091–1102 (2019).

27. Trubetskoy, V. et al. Mapping genomic loci implicates genes and synaptic biology in schizophrenia. Nature 604, 502–508 (2022).

28. International Multiple Sclerosis Genetics Consortium. Multiple sclerosis genomic map implicates peripheral immune cells and microglia in susceptibility. Science 365, eaav7188 (2019).

29. van Rheenen, W. et al. Common and rare variant association analyses in amyotrophic lateral sclerosis identify 15 risk loci with distinct genetic architectures and neuron-specific biology. Nat. Genet. 53, 1636–1648 (2021).

30. Paolicelli, R. C. et al. Microglia states and nomenclature: A field at its crossroads. Neuron 110, 3458–3483 (2022).

31. Sun, N. et al. Human microglial state dynamics in Alzheimer’s disease progression. Cell 186, 4386–4403.e29 (2023).

32. Tuddenham, J. F. et al. A cross-disease resource of living human microglia identifies disease-enriched subsets and tool compounds recapitulating microglial states. Nat. Neurosci. 27, 2521–2537 (2024).

33. Thrupp, N. et al. Single-nucleus RNA-seq is not suitable for detection of microglial activation genes in humans. Cell Rep. 32, 108189 (2020).

34. The Synapse Engineering Team. Synapseclient: A Client for Synapse, a Collaborative Compute Space That Allows Scientists to Share and Analyze Data Together.

35. https://github.com/hail-is/hail/releases/tag/0.2.13.

36. Pedersen, B. S. et al. Somalier: rapid relatedness estimation for cancer and germline studies using efficient genome sketches. Genome Med. 12, 62 (2020).

37. Wolf, F. A., Angerer, P. & Theis, F. J. SCANPY: large-scale single-cell gene expression data analysis. Genome Biol. 19, 15 (2018).

38. Wolock, S. L., Lopez, R. & Klein, A. M. Scrublet: Computational identification of cell Doublets in Single-cell transcriptomic data. Cell Syst. 8, 281–291.e9 (2019).

39. Korsunsky, I. et al. Fast, sensitive and accurate integration of single-cell data with Harmony. Nat. Methods 16, 1289–1296 (2019).

40. Stegle, O., Parts, L., Durbin, R. & Winn, J. A Bayesian framework to account for complex non-genetic factors in gene expression levels greatly increases power in eQTL studies. PLoS Comput. Biol. 6, e1000770 (2010).

41. GTEx Consortium et al. Genetic effects on gene expression across human tissues. Nature 550, 204–213 (2017).

42. Yang, J., Lee, S. H., Goddard, M. E. & Visscher, P. M. GCTA: a tool for genome-wide complex trait analysis. Am. J. Hum. Genet. 88, 76–82 (2011).

43. Purcell, S. et al. PLINK: a tool set for whole-genome association and population-based linkage analyses. Am. J. Hum. Genet. 81, 559–575 (2007).

44. Sapiro, A. L. et al. Zinc finger RNA-binding protein Zn72D regulates ADAR-mediated RNA editing in neurons. Cell Rep. 31, 107654 (2020).

45. Tang, Y., Ghosal, S. & Roy, A. Nonparametric bayesian estimation of positive false discovery rates. Biometrics 63, 1126–1134 (2007).

46. Mullins, N. et al. Genome-wide association study of more than 40,000 bipolar disorder cases provides new insights into the underlying biology. Nat. Genet. 53, 817–829 (2021).

47. Collins, R. L. et al. A structural variation reference for medical and population genetics. Nature 581, 444–451 (2020).

48. Myers, T. A., Chanock, S. J. & Machiela, M. J. LDlinkR: An R package for rapidly calculating linkage disequilibrium statistics in diverse populations. Front. Genet. 11, 157 (2020).

49. Hemani, G. et al. The MR-Base platform supports systematic causal inference across the human phenome. Elife 7, (2018).

50. 1000 Genomes Project Consortium et al. A global reference for human genetic variation. Nature 526, 68–74 (2015).

51. Hartwig, F. P., Davey Smith, G. & Bowden, J. Robust inference in summary data Mendelian randomization via the zero modal pleiotropy assumption. Int. J. Epidemiol. 46, 1985–1998 (2017).

52. Bowden, J., Davey Smith, G., Haycock, P. C. & Burgess, S. Consistent estimation in Mendelian randomization with some invalid instruments using a weighted median estimator. Genet. Epidemiol. 40, 304–314 (2016).

53. Bowden, J., Davey Smith, G. & Burgess, S. Mendelian randomization with invalid instruments: effect estimation and bias detection through Egger regression. Int. J. Epidemiol. 44, 512–525 (2015).

54. Cooler, S. & Schwartz, G. W. An offset ON-OFF receptive field is created by gap junctions between distinct types of retinal ganglion cells. Nat. Neurosci. 24, 105–115 (2021).

55. Weissbrod, O. et al. Functionally informed fine-mapping and polygenic localization of complex trait heritability. Nat. Genet. 52, 1355–1363 (2020).

56. Fang, R. et al. Mapping of long-range chromatin interactions by proximity ligation-assisted ChIP-seq. Cell Res. 26, 1345–1348 (2016).

57. Lawrence, M. et al. Software for computing and annotating genomic ranges. PLoS Comput. Biol. 9, e1003118 (2013).

