## Supplementary Note for "SingleBrain: A Meta-Analysis of Single-Nucleus eQTLs Linking Genetic Risk to Brain Disorders"

**Table of Contents**

**Supplementary Methods** **3**
Genotype data quality control 3
Visualization 3
**Supplementary Figures** **4**
Supplementary Figure 1 - Demographics of the SingleBrain cohorts. 5
Supplementary Figure 2 - Principal component analysis of SingleBrain participants projected on reference populations from the 1000 Genomes Project samples. 5
Supplementary Figure 3 - eGene discovery in each cell type and subtype. 6
Supplementary Figure 4 - Chromatin states of brain cell type eQTLs. 7
Supplementary Figure 5 - Pairwise sharing of genes with significant lead eQTLs from total cell class compared to reference brain eQTLs. 8
Supplementary Figure 6 - Pairwise sharing of genes with significant lead eQTLs from total cell class. 9
Supplementary Figure 7 - Estimated proportion of 6 neurological disease heritability mediated by brain major cell types cis-eQTL. 10
Supplementary Figure 8 - All SCZ GWAS loci with PP4 > 0.8 and significant MR in SingleBrain and MiGA. 11-12
Supplementary Figure 9 - All ALS, BDP, and MS GWAS loci with PP4 > 0.8 and significant MR in SingleBrain and MiGA. 13
Supplementary Figure 10 - Analysis of the *PBX1* excitatory neuron eQTL with BPD GWAS *NUF2* locus. 14-15
Supplementary Figure 11 - Number of GWAS loci with at least one colocalization (probability of sharing one causal variant between eQTL and GWAS [PP4] > 0.8) and fine-mapping loci (PIP > 0.95). 16
Supplementary Figure 12 - Volcano plot of SCZ MPRA with SNPs mapped to the *DPP* SCZ GWAS locus. 17
Supplementary Figure 13 - Number of SCZ GWAS loci with a PP4 > 0.8 and excitatory neurons subtype colocalization. 18-19
Supplementary Figure 14 - Number of AD and PD GWAS loci with PP4 > 0.8. 20
Supplementary Figure 15 - All 3 AD GWAS loci with PP4 > 0.8 and significant MR in SingleBrain and MiGA. 21
Supplementary Figure 16 - All AD and PD GWAS locus with PP4 > 0.8 colocalization at SingleBrain microglia subtypes and MiGA. 22
Supplementary Figure 17 - Locus zoom and fine-mapping of *COX7C* and *SORL1* AD GWAS locus with SingleBrain MG and MiGA eQTL. 23
Supplementary Figure 18 - Comparison of the maximum SingleBrain microglia subtypes and MiGA PP4 for each PD GWAS locus across both references. 24
Supplementary Figure 19 - Overlap SNPs dot plot compared to *FGF20* PD GWAS loci with *FGF20* eQTLs. 25
Supplementary Figure 20 - Analysis of the *FGF20* MG3 eQTL with PD GWAS *FGF20* locus. 26-27

### Supplementary Methods

##### Genotype data quality control

To perform the whole-genome sequencing (WGS) data quality control (QC), we used the Python library Hail[1] (v.0.2.113). We split multiallelic variants into biallelic variants. Variants with low Variant Quality Score Recalibration quality or that resided within known low-complexity regions were filtered out. Next, genotype QC was performed. Sample-level QC removed samples with a low mean depth (< 0.20), a low call rate (< 0.99), and mismatches between the genotype and sex. We excluded non-European samples estimated by Somalier[2] (v.0.2.12). In the European population samples, outliers with a variant count outside 4 or 8 standard deviations (SDs) from the mean were removed. After removing samples that did not pass the sample-level QC, we performed the variant-level QC with the following criteria: monomorphic variants (allele count [AC] = 0), low variant call rate (< 0.90), those that failed the Hardy-Weinberg equilibrium test with the unrelated European samples ($P < 10^{-9}$), low variant quality scores. For single-nucleotide polymorphisms (SNPs), variants with QD ≥ 2, SOR ≤ 3, FS ≤ 60, MQ ≤ 40, MQRankSum ≤ |12.5|, and ReadPosRankSum ≤ |12.5| were removed. For insertions and deletions (INDELs), variants with QD ≥ 2, FS ≤ 200, and ReadPosRankSum ≤ |20| were removed. We annotated the RSID using bcftools[3] v.1.9 and retained common variants (allele frequency [AF] > 0.01).

Abbreviations: QD, quality by depth; SOR, strand odds ratio; FS, Fisher strand; MQ, root mean square mapping quality; MQRankSum, rank sum test results for mapping quality; ReadPosRankSum, rank sum test results for site position within reads

##### Visualization

All plots were created using ggplot2 (version 3.3.3) in R (v4.0.4), with ggrepel (v.0.9.1) for additional layers of visualization.

### Supplementary Figures

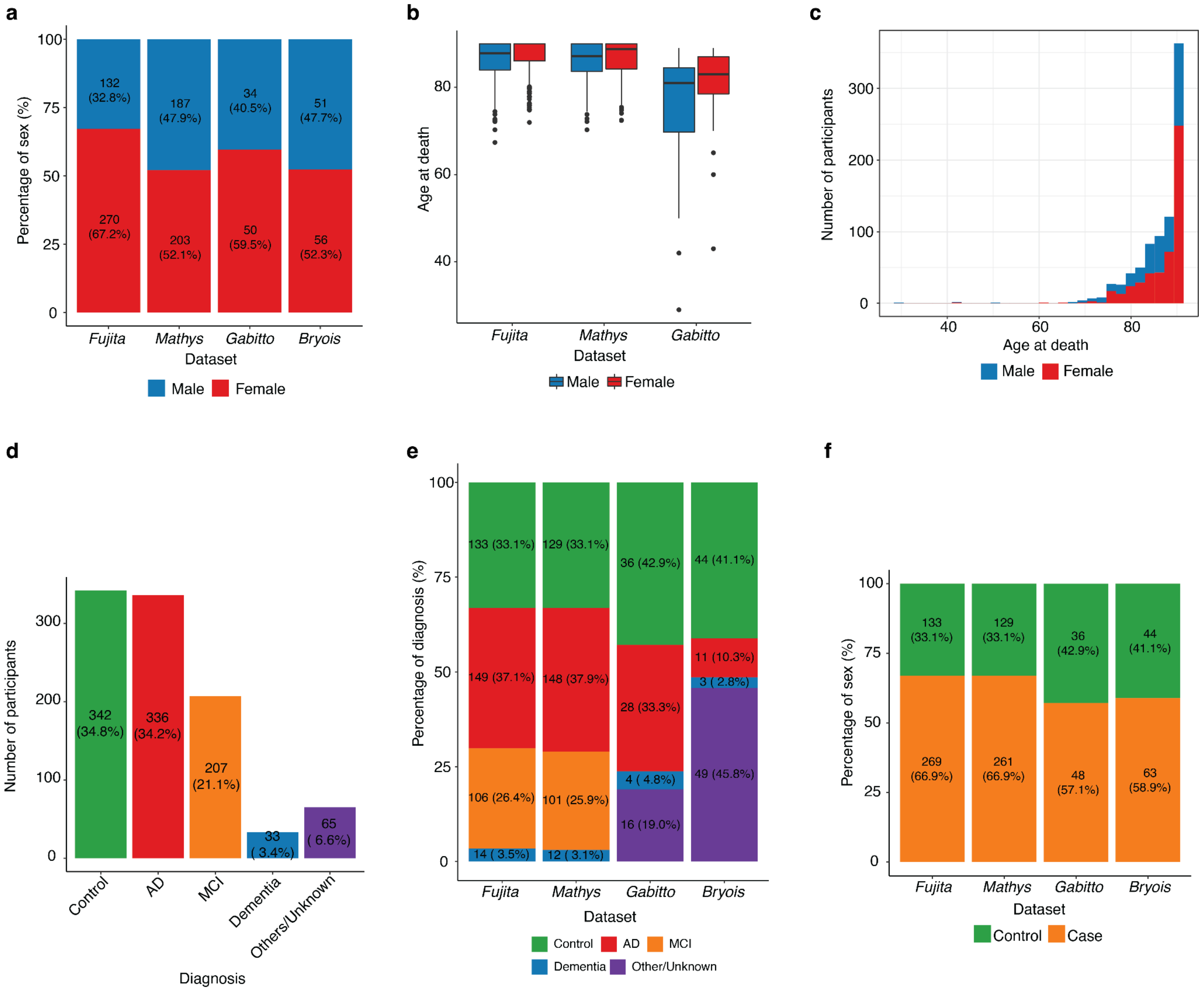


**Supplementary Figure 1 - Demographics of the SingleBrain cohorts.**

**a)** Proportions of donors by sex in each cohort. **b-c)** Age range of the donors in each cohort. Blue indicates the distribution of male donors and red indicates the distribution of female donors. **d)** Total number of diagnoses in this study. **e)** Proportions of diagnoses in each cohort. **f)** Number of cases and controls in each cohort.

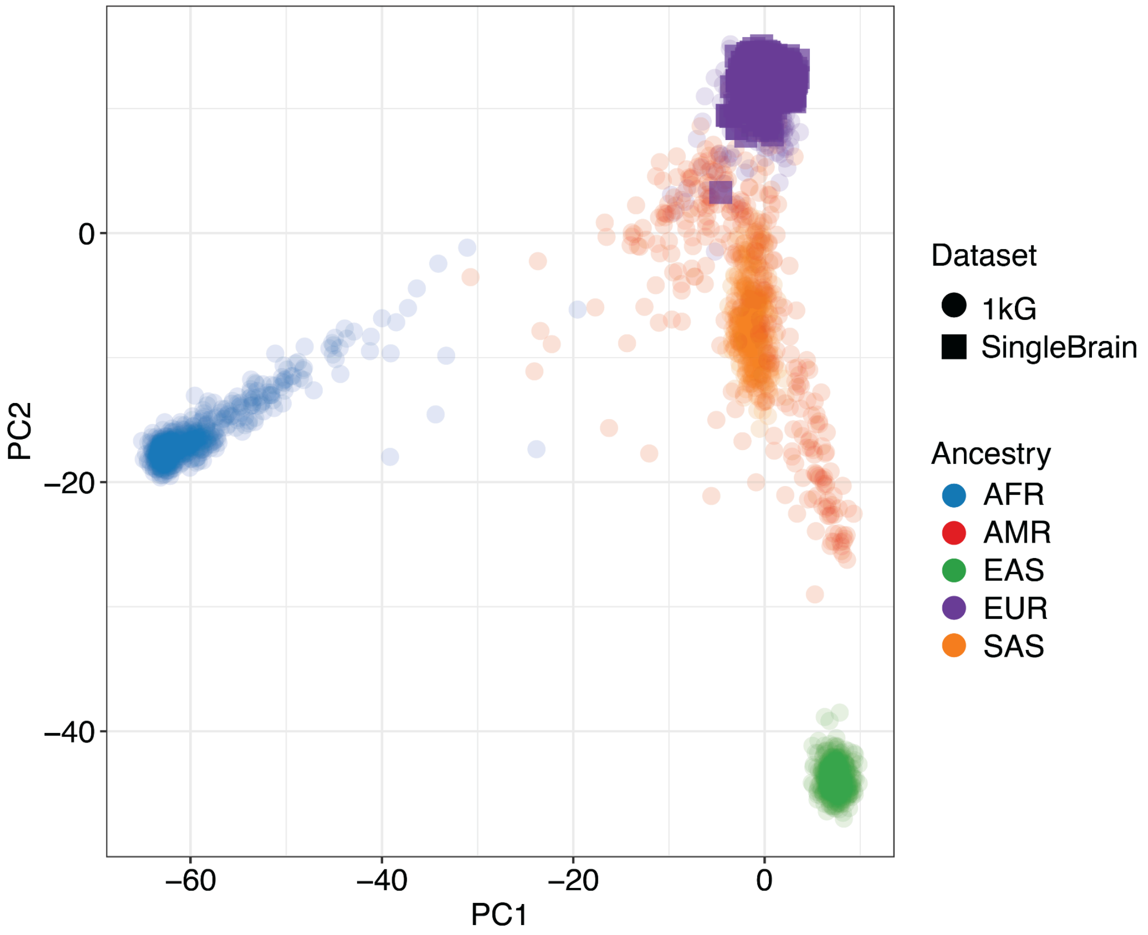


**Supplementary Figure 2 - Principal component analysis (PCA) of SingleBrain participants projected on reference populations from the 1000 Genomes Project samples.**

PCA was performed to project SingleBrain study participants onto a reference space derived from the 1000 Genomes Project samples. The 983 participants from the SingleBrain dataset (represented by purple squares) were mapped onto the PCA space to assess their ancestral background relative to 1000 Genomes Populations. PCA, principal component analysis; PC, principal component; 1kG, 1000 Genomes Project; AFR, African; AMR, American; EAS, East Asian; EUR, European; SAS, South Asian.

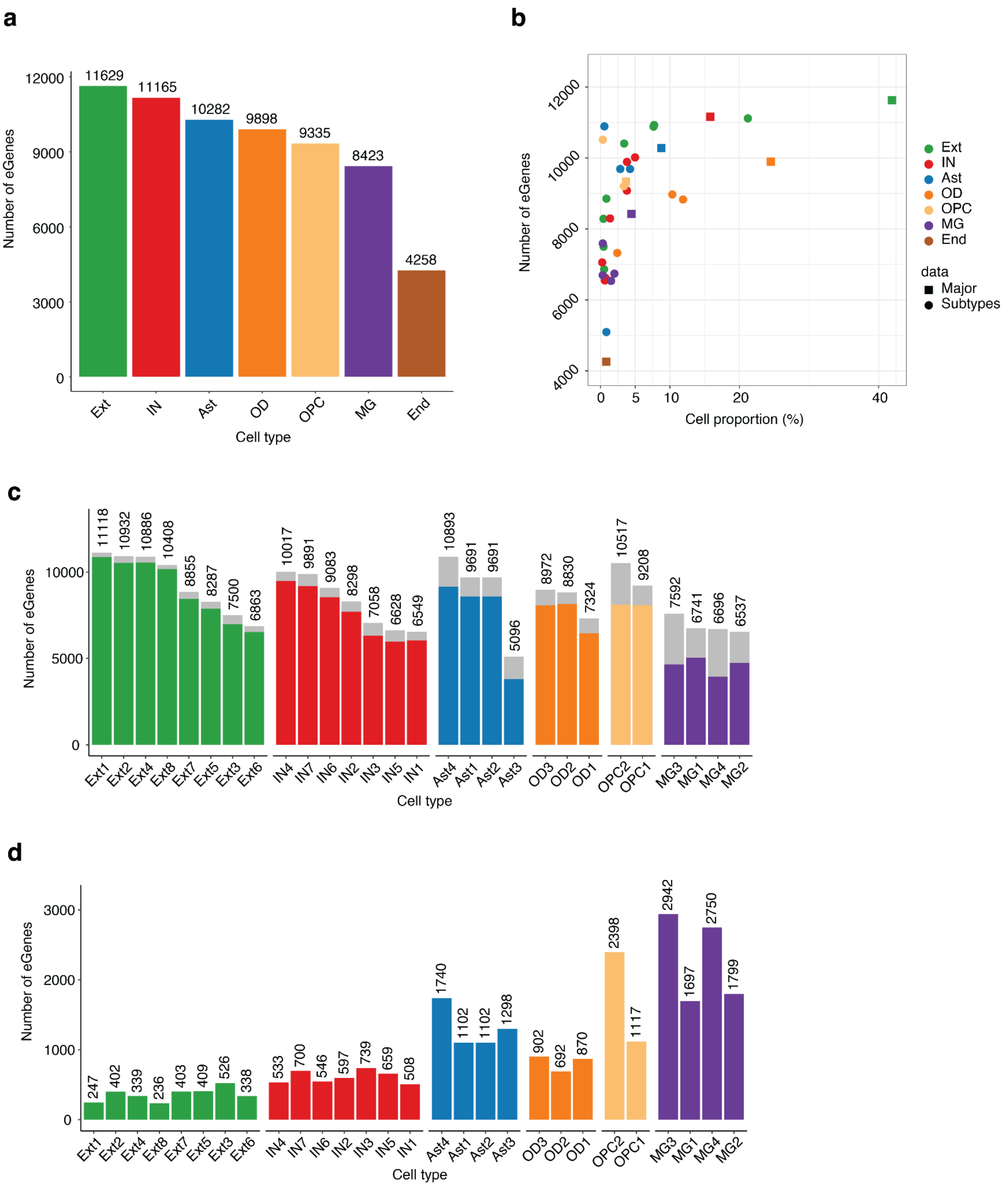


**Supplementary Figure 3 - eGene discovery in each cell type and subtype.**

**a)** Number of eGenes detected within the brain's seven cell types. **b)** Number of eGenes discovered plotted against each cell's proportion of the 35 cell types in this study. Squares indicate the brain's seven major cell types. Triangles indicate subtypes. **c)** Number of eGenes detected in each of the brain cell subtypes. The gray color indicates the number of eGenes unique to each subtype. **d)** Number of eGenes specific to each cell subtype. Ext, excitatory neurons; IN, inhibitory neurons; OD, oligodendrocytes; Ast, astrocytes; OPC, oligodendrocyte progenitor cells; MG, microglia; End, endothelial cells.

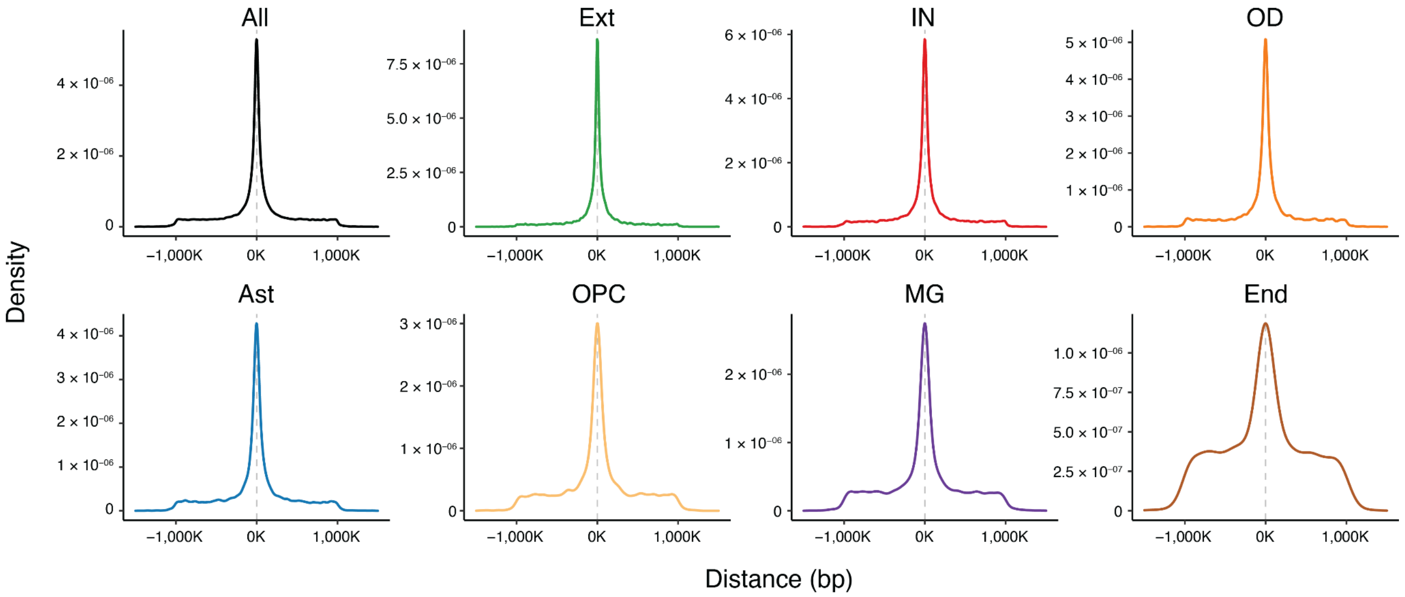


**Supplementary Figure 4 - Chromatin states of brain cell type expression quantitative trait loci (eQTLs).**

Density of distance between eQTL lead SNPs and transcription start site (TSS). Ext, excitatory neurons; IN, inhibitory neurons; OD, oligodendrocytes; Ast, astrocytes; OPC, oligodendrocyte progenitor cells; MG, microglia; End, endothelial cells.

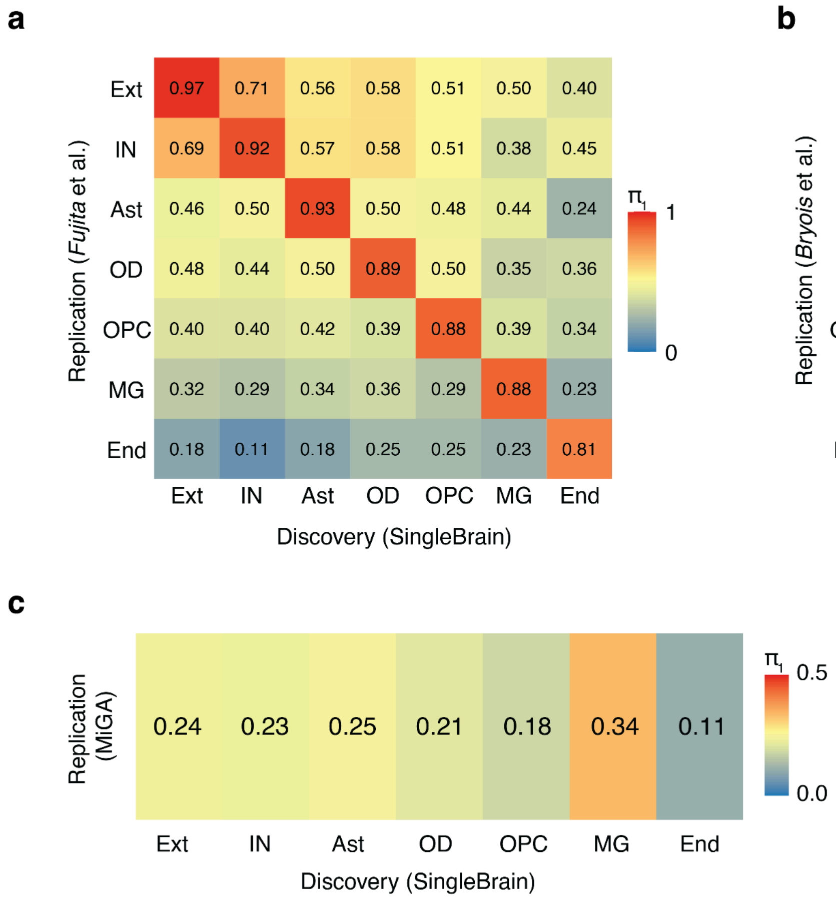


**Supplementary Figure 5 - Pairwise sharing of genes with significant lead eQTLs from total cell class compared to reference brain eQTLs.**

The current dataset (columns) is used for eQTL discovery, and the replication rate is **a)** *Fujita* et al.[4], **b)** *Bryois* et al.[5] and **c)** MiGA[6], respectively. The analysis was based on Storey's $\pi_1$. Rates of sharing ($\pi_1$: the proportion of true alternative hypotheses) between two QTL results were estimated by leveraging the distribution of p-values and adjusting the false discovery rate (FDR) through the q-value framework. MiGA; Microglia Genomic Atlas, Ext, excitatory neurons; IN, inhibitory neurons; OD, oligodendrocytes; Ast, astrocytes; OPC, oligodendrocyte progenitor cells; MG, microglia; End, endothelial cells.

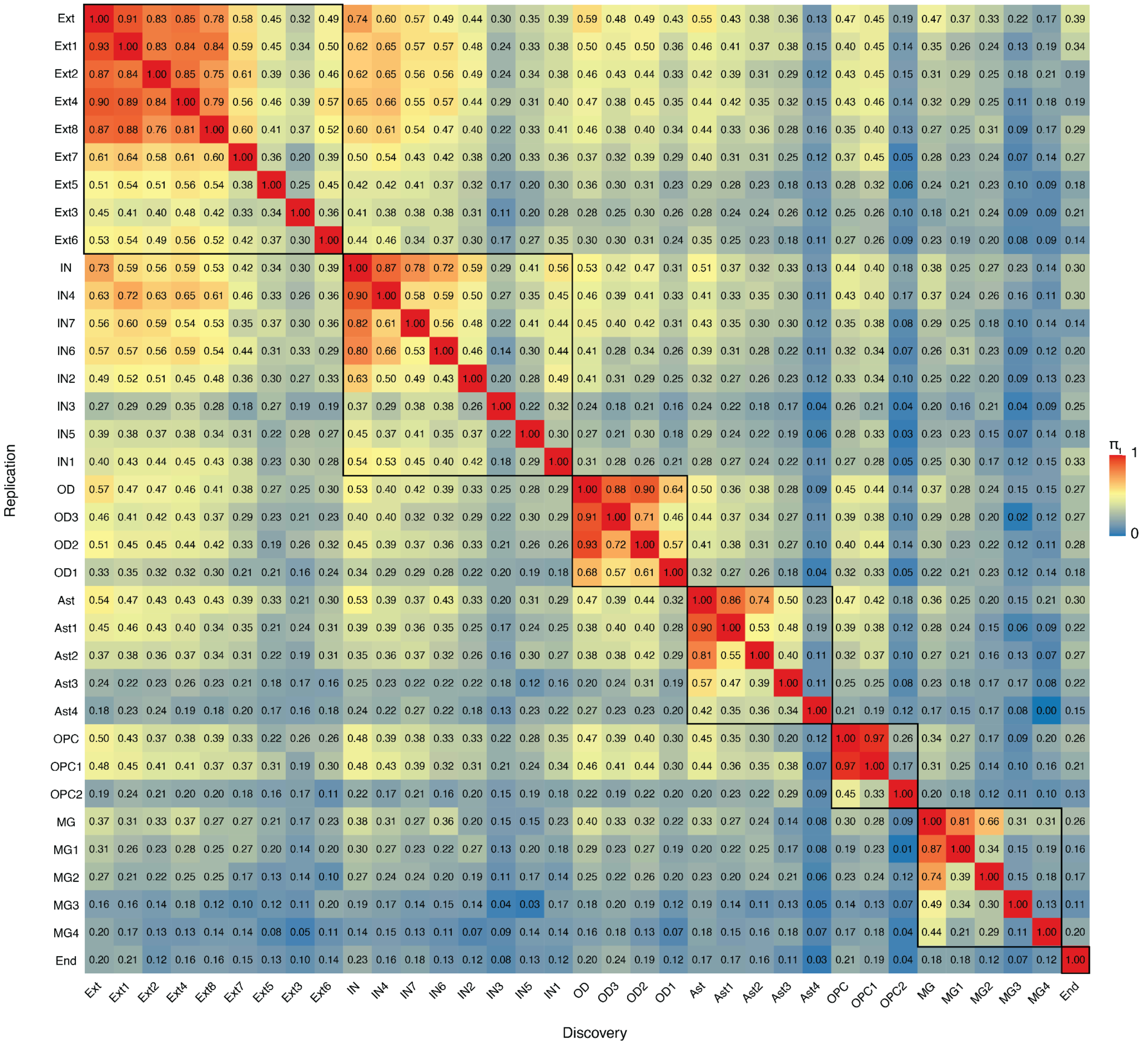


**Supplementary Figure 6 - Pairwise sharing of genes with significant lead eQTLs from total cell class.**

The analysis was based on Storey's $\pi_1$. Rates of sharing ($\pi_1$: the proportion of true alternative hypotheses) between two QTL results were estimated by leveraging the distribution of p-values and adjusting the false discovery rate (FDR) through the q-value framework. The columns are used for brain eQTL lead SNPs of this study, and the replication rate is each cell type QTLs in discovery SNPs, respectively. Ext, excitatory neurons; IN, inhibitory neurons; OD, oligodendrocytes; Ast, astrocytes; OPC, oligodendrocyte progenitor cells; MG, microglia; End, endothelial cells.

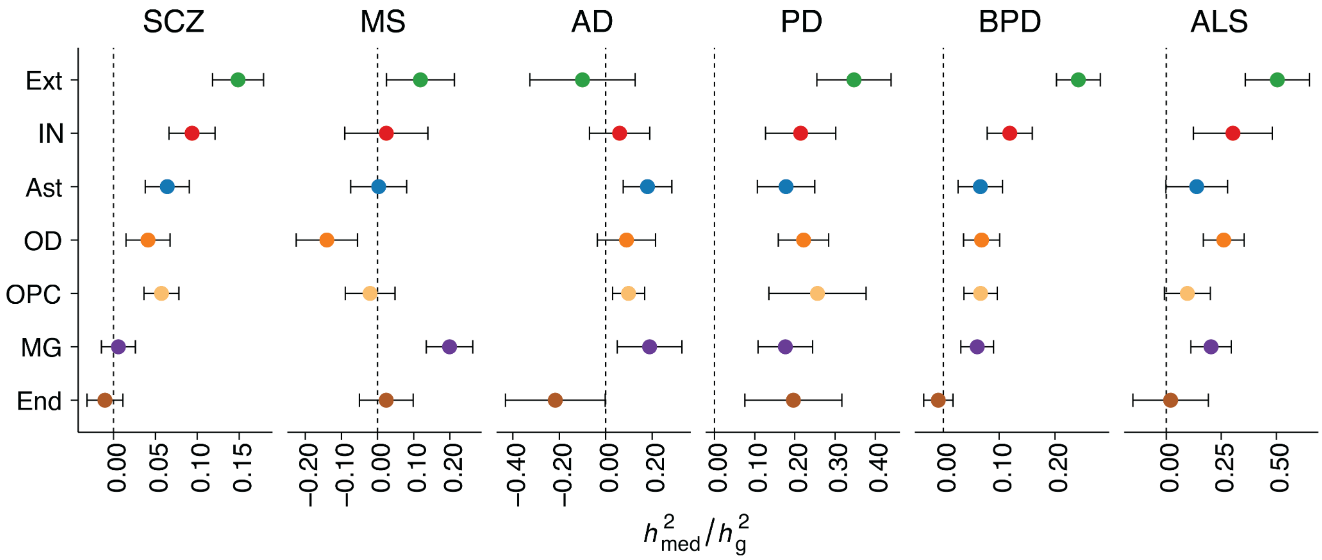


**Supplementary Figure 7 - Estimated proportion of 6 neurological disease heritability mediated by brain major cell types *cis*-eQTL.** Disease heritability ($h^2_{med}/h^2_g$) using Mediated Expression Score Regression (MESC). MESC estimates the proportion of heritability mediated by the *cis*-genetic component. SCZ, schizophrenia; MS, multiple sclerosis; AD, Alzheimer's disease; PD, Parkinson's disease; BPD, bipolar disorder; ALS, amyotrophic lateral sclerosis; Ext, excitatory neurons; IN, inhibitory neurons; OD, oligodendrocytes; Ast, astrocytes; OPC, oligodendrocyte progenitor cells; MG, microglia; End, endothelial cells.

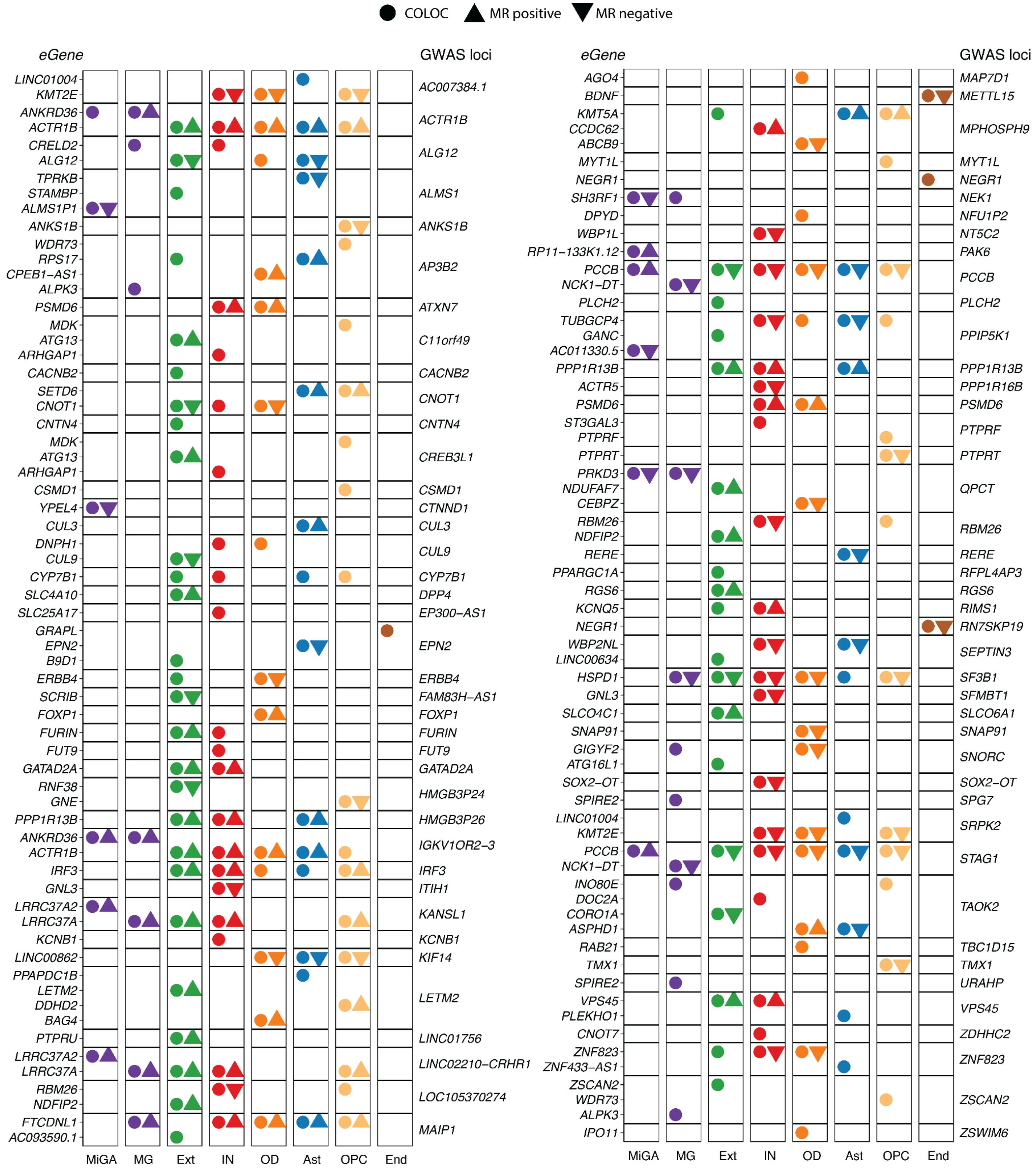


**Supplementary Figure 8 - All SCZ GWAS loci with PP4 > 0.8 and significant MR association in SingleBrain and MiGA.**

The circle represents eQTL with PP4 > 0.8. The triangle denotes significant genes in MR results, with the orientation of the triangle corresponding to the direction of effect. COLOC, colocalization; MR, Mendelian randomization; MiGA, Microglia Genomic Atlas; MG, microglia; Ext, excitatory neurons; IN, inhibitory neurons; Ast, astrocytes; OD, oligodendrocytes; OPC, oligodendrocyte progenitor cells; End, endothelial cells.

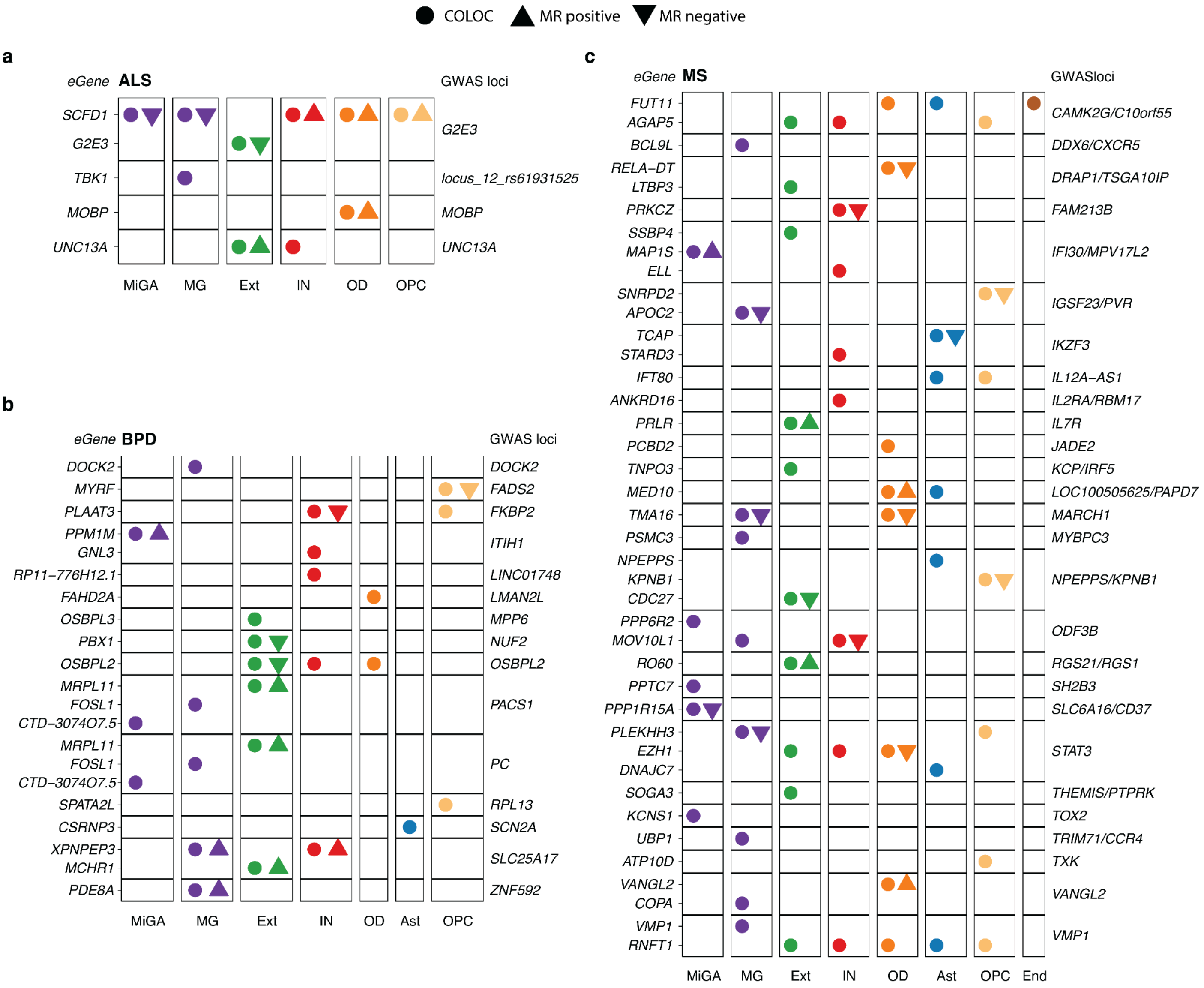


**Supplementary Figure 9 - All ALS, BDP, and MS GWAS loci with PP4 > 0.8 and significant MR association in SingleBrain and MiGA.**

The circle represents eQTL with PP4 > 0.8. The triangle denotes significant genes in MR results, with the orientation of the triangle corresponding to the direction of effect. **a)** Amyotrophic lateral sclerosis (ALS). **b)** Bipolar disease (BPD). **c)** Multiple sclerosis (MS). ALS, amyotrophic lateral sclerosis; BPD, bipolar disease; MS, multiple sclerosis; COLOC, colocalization; MR, Mendelian randomization; MiGA, Microglia Genomic Atlas; MG, microglia; Ext, excitatory neurons; IN, inhibitory neurons; Ast, astrocytes; OD, oligodendrocytes; OPC, oligodendrocyte progenitor cells; End, endothelial cells.

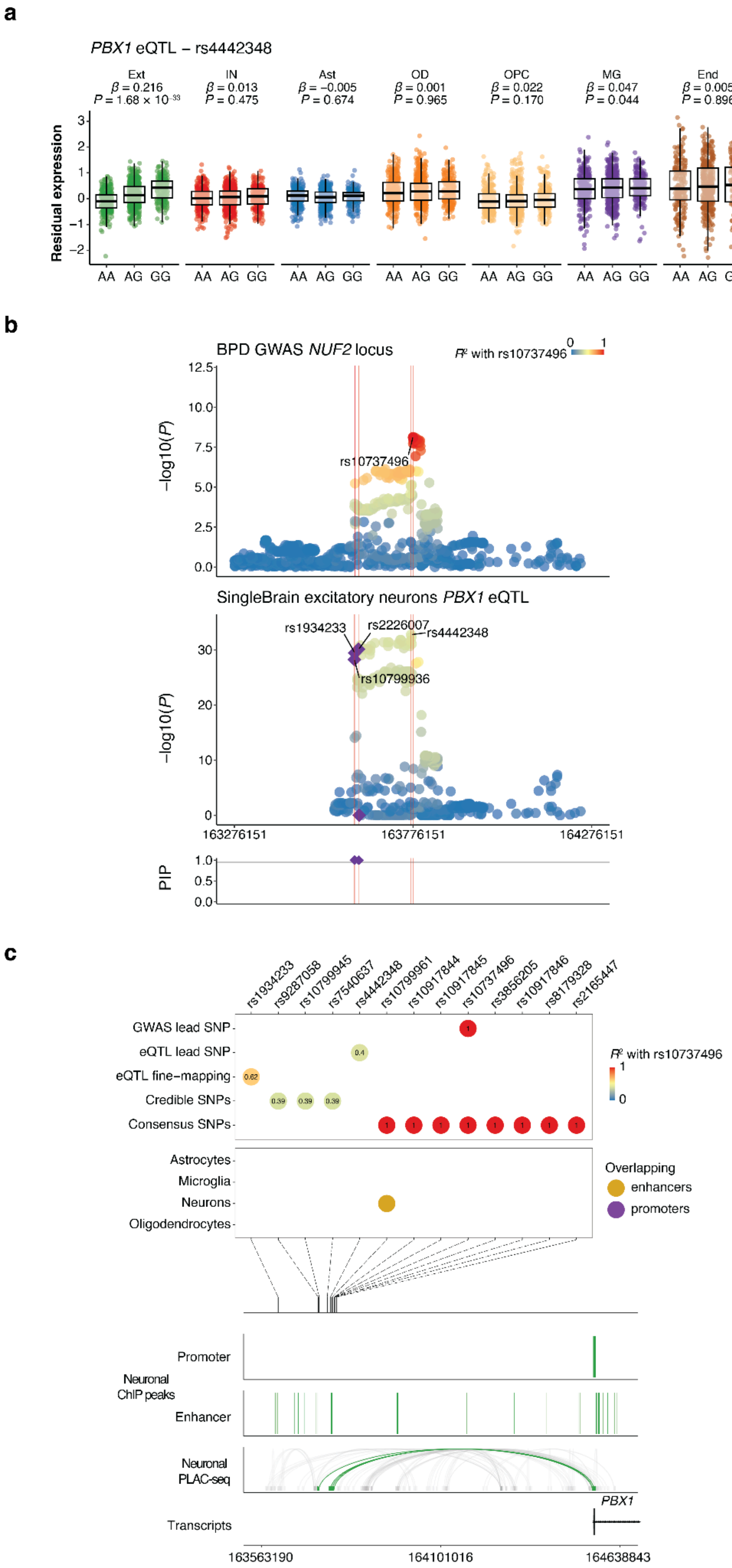
a
PBX1 eQTL – rs4442348
Ext
β = 0.216
P = 1.68 × 10⁻³³
IN
β = 0.013
P = 0.475
Ast
β = −0.005
P = 0.674
OD
β = 0.001
P = 0.965
OPC
β = 0.022
P = 0.170
MG
β = 0.047
P = 0.044
End
β = 0.005
P = 0.896
Residual expression
AA AG GG
b
BPD GWAS NUF2 locus
R² with rs10737496
rs10737496
−log10(P)
SingleBrain excitatory neurons PBX1 eQTL
rs1934233
rs2226007
rs4442348
rs10799936
163276151
163776151
164276151
PIP
c
rs1934233
rs9287058
rs10799945
rs7540637
rs4442348
rs10799961
rs10917844
rs10917845
rs10737496
rs3856205
rs10917846
rs8179328
rs2165447
GWAS lead SNP
eQTL lead SNP
eQTL fine−mapping
Credible SNPs
Consensus SNPs
R² with rs10737496
Astrocytes
Microglia
Neurons
Oligodendrocytes
Overlapping
enhancers
promoters
Promoter
Neuronal ChIP peaks
Enhancer
Neuronal PLAC-seq
PBX1
Transcripts
163563190
164101016
164638843

**Supplementary Figures 10 - Analysis of the *PBX1* excitatory neuron eQTL with BPD GWAS *NUF2* locus.**

**a)** *PBX1* expression is associated with the rs4442348 genotype, specifically in excitatory neurons. The residual expression is PEER adjusted gene expression level. The nominal *P*-value and beta from the linear regression model in the brain cell type eQTL analysis are indicated above the box plots. The box plots show the median, the box spans from the first to the third quartiles, and the whiskers extend 1.5 times the interquartile range (IQR) from the box. **b)** Locus zoom and fine-mapping of the *NUF2* BPD GWAS and *PBX1* excitatory neurons eQTL. Labels refer to lead SNPs and fine-mapping SNPs with $P < 1 \times 10^{-4}$. SNPs are colored by the LD with the lead GWAS SNP. Fine-mapped SNPs with PIP > 0.95 from GWAS and eQTL are presented under the plot. **c)** Fine-mapping of the *NUF2* locus and combination with the GWAS lead SNP, fine-mapping SNPs, eQTL lead SNP, fine-mapping SNPs, and credible SNPs of colocalization. SNPs are colored by the LD with the lead GWAS SNP, overlapped with cell-type-specific enhancers and promoters were defined by *Nott* et al.[7]. Genomic plots (hg19) of the lead SNPs and fine-mapping SNPs with $P < 1 \times 10^{-4}$ and epigenomic data from the microglia ChIP-seq and PLAC-seq junctions. BPD, bipolar disease; Ext, excitatory neurons; IN, inhibitory neurons; OD, oligodendrocytes; Ast, astrocytes; OPC, oligodendrocyte progenitor cells; MG, microglia; End, endothelial cells.

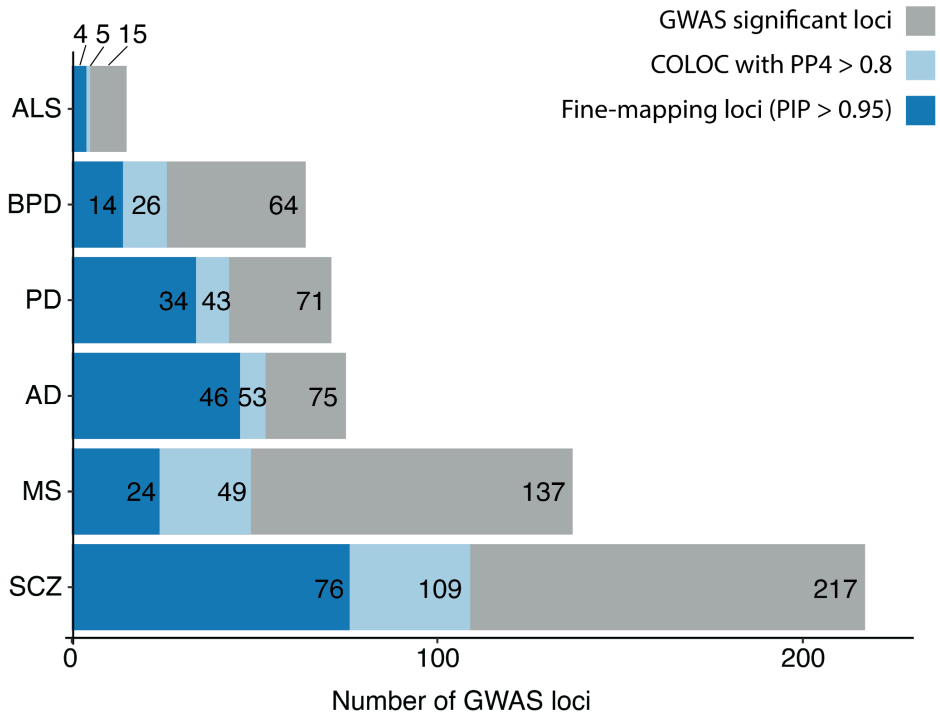


**Supplementary Figure 11 - Number of GWAS loci with at least one colocalization (probability of sharing one causal variant between eQTL and GWAS [PP4] > 0.8) and fine-mapping loci (PIP > 0.95).**

The dark blue indicates consistent loci between the COLOC and fine-mapping loci (PIP > 0.95). The light blue indicates the only COLOC locus. The gray indicates loci with no COLOC evidence (PP4 < 0.8). ALS, amyotrophic lateral sclerosis; AD, Alzheimer's disease; BPD, bipolar disorder; PD, Parkinson's disease; AD, Alzheimer's disease; MS, multiple sclerosis; SCZ, schizophrenia.

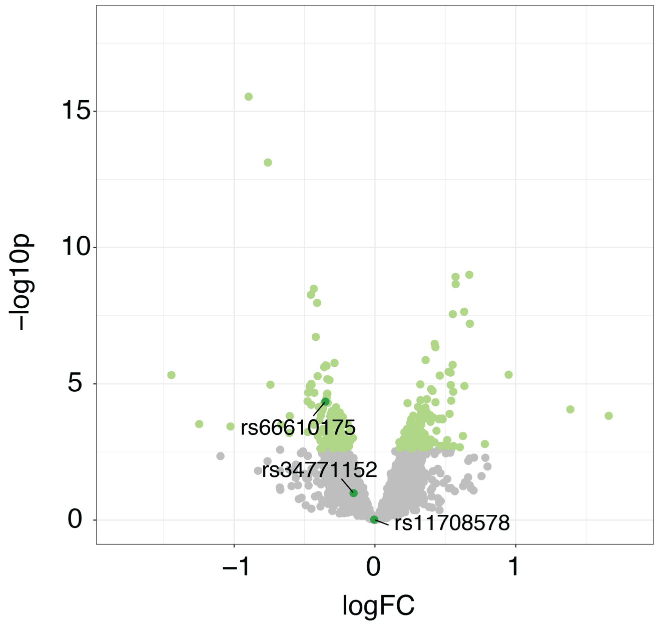


**Supplementary Figure 12 - Volcano plot of SCZ MPRA with SNPs mapped to the *DPP* SCZ GWAS locus[8].**

The analysis was based on the linear model test. Using the paired mixed-model fit, differential regulatory activity was measured in the DNA/RNA matrix between two alleles through multiple tests using the Bonferroni correction. The gray color indicates variants, and the light green color indicates FDR < 0.05. The dark green dot indicates the *DDP* SCZ GWAS locus variants.

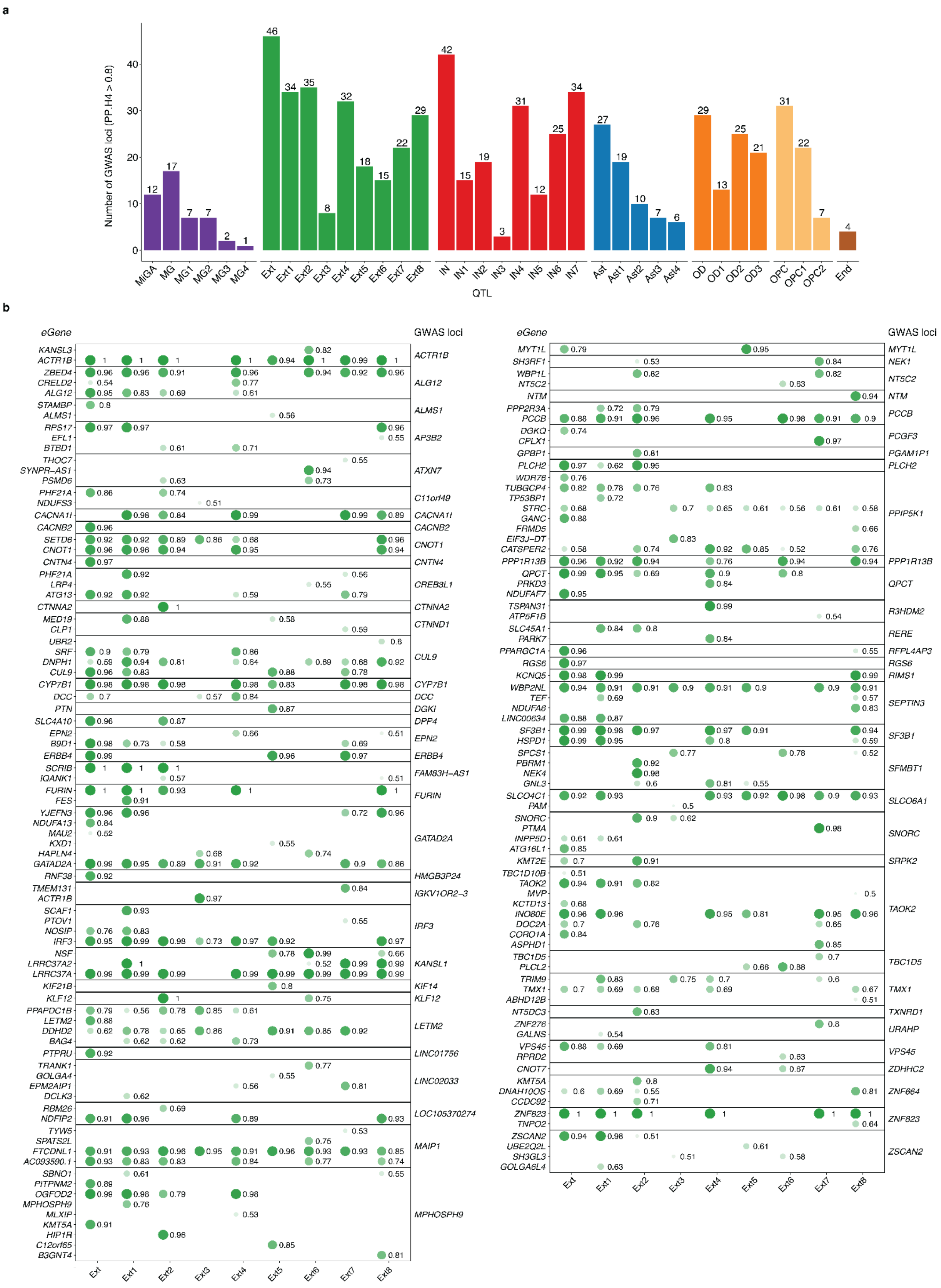

a
Number of GWAS loci (PP.H4 > 0.8)
12 17 7 7 2 1
46 34 35 8 32 18 15 22 29
42 15 19 3 31 12 25 34
27 19 10 7 6
29 13 25 21
31 22 7
4
MiGA MG MG1 MG2 MG3 MG4 Ext Ext1 Ext2 Ext3 Ext4 Ext5 Ext6 Ext7 Ext8 IN IN1 IN2 IN3 IN4 IN5 IN6 IN7 Ast Ast1 Ast2 Ast3 Ast4 OD OD1 OD2 OD3 OPC OPC1 OPC2 End
QTL
b
eGene
GWAS loci
ACTR1B ALG12 ALMS1 AP3B2 ATXN7 C11orf49 CACNA1I CACNB2 CNOT1 CNTN4 CREB3L1 CTNNA2 CTNND1 CUL9 CYP7B1 DCC DGKI DPP4 EPN2 ERBB4 FAM83H-AS1 FURIN GATAD2A HMGB3P24 IGKV1OR2-3 IRF3 KANSL1 KIF14 KLF12 LETM2 LINC01756 LINC02033 LOC105370274 MAIP1 MPHOSPH9
MYT1L NEK1 NT5C2 NTM PCCB PCGF3 PGAM1P1 PLCH2 PPIP5K1 PPP1R13B QPCT R3HDM2 RERE RFPL4AP3 RGS6 RIMS1 SEPTIN3 SF3B1 SFMBT1 SLCO6A1 SNORC SRPK2 TAOK2 TBC1D5 TMX1 TXNRD1 URAHP VPS45 ZDHHC2 ZNF664 ZNF823 ZSCAN2
Ext Ext1 Ext2 Ext3 Ext4 Ext5 Ext6 Ext7 Ext8

**Supplementary Figure 13 - Number of SCZ GWAS loci with a PP4 > 0.8 and excitatory neurons subtype colocalization.**

**a)** Number of SCZ GWAS loci (PP4 > 0.8). **b)** All SCZ GWAS locus had a PP4 > 0.8 colocalization at the excitatory neurons subtype. Shape opacity and size were scaled to the magnitude of PP4. The numbers next to the circles are PP4 values. MiGA, Microglia Genomic Atlas; MG, microglia; Ext, excitatory neurons; IN, inhibitory neurons; Ast, astrocytes; OD, oligodendrocytes; OPC, oligodendrocyte progenitor cells; End, endothelial cells.

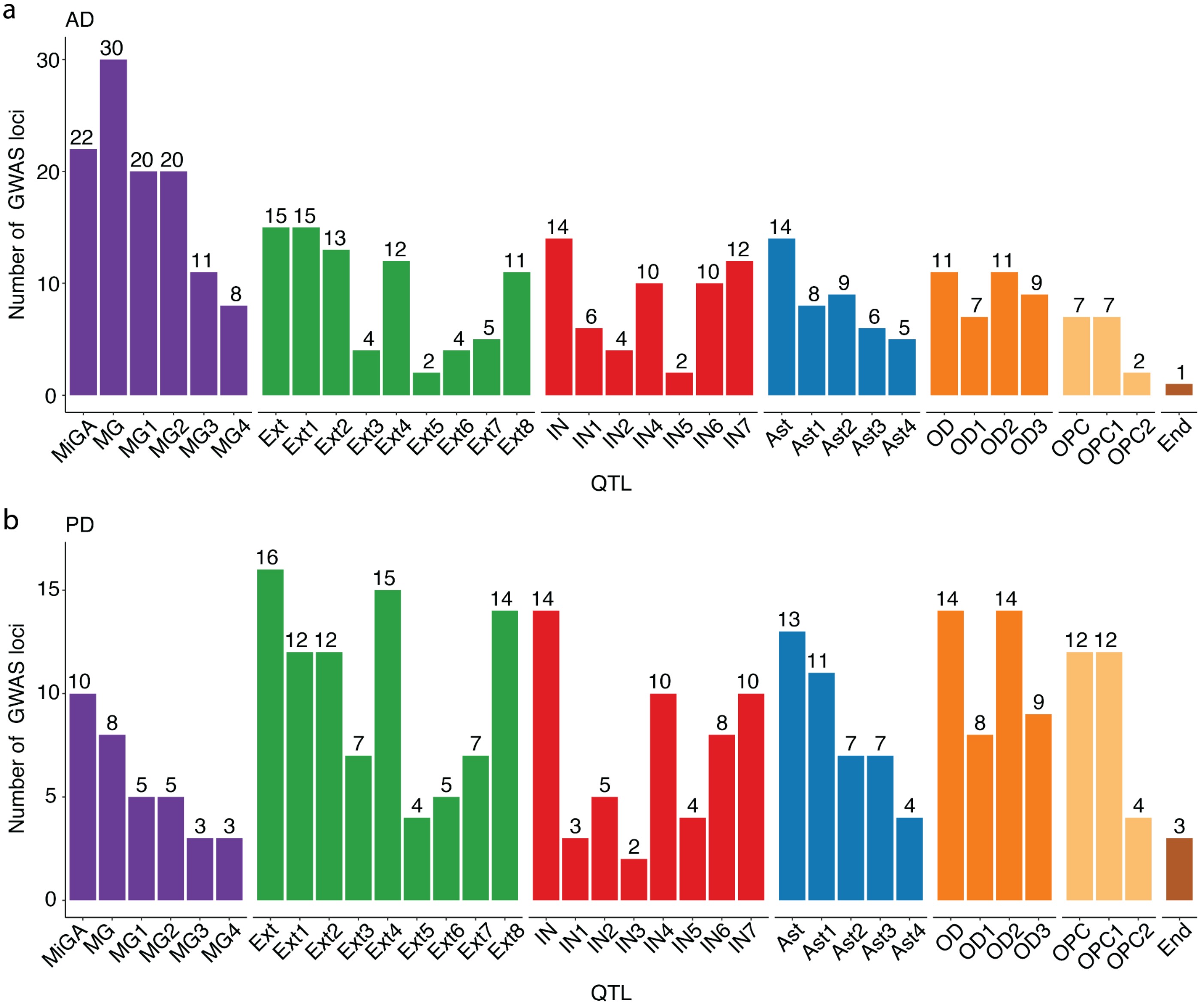


**Supplementary Figure 14 - Number of AD and PD GWAS loci with PP4 > 0.8.**

**a)** Alzheimer's disease. **b)** Parkinson's disease. AD, Alzheimer's disease; PD, Parkinson's disease; MiGA, Microglia Genomic Atlas; MG, microglia; Ext, excitatory neurons; IN, inhibitory neurons; OD, oligodendrocytes; Ast, astrocytes; OPC, oligodendrocyte progenitor cells; End, endothelial cells.

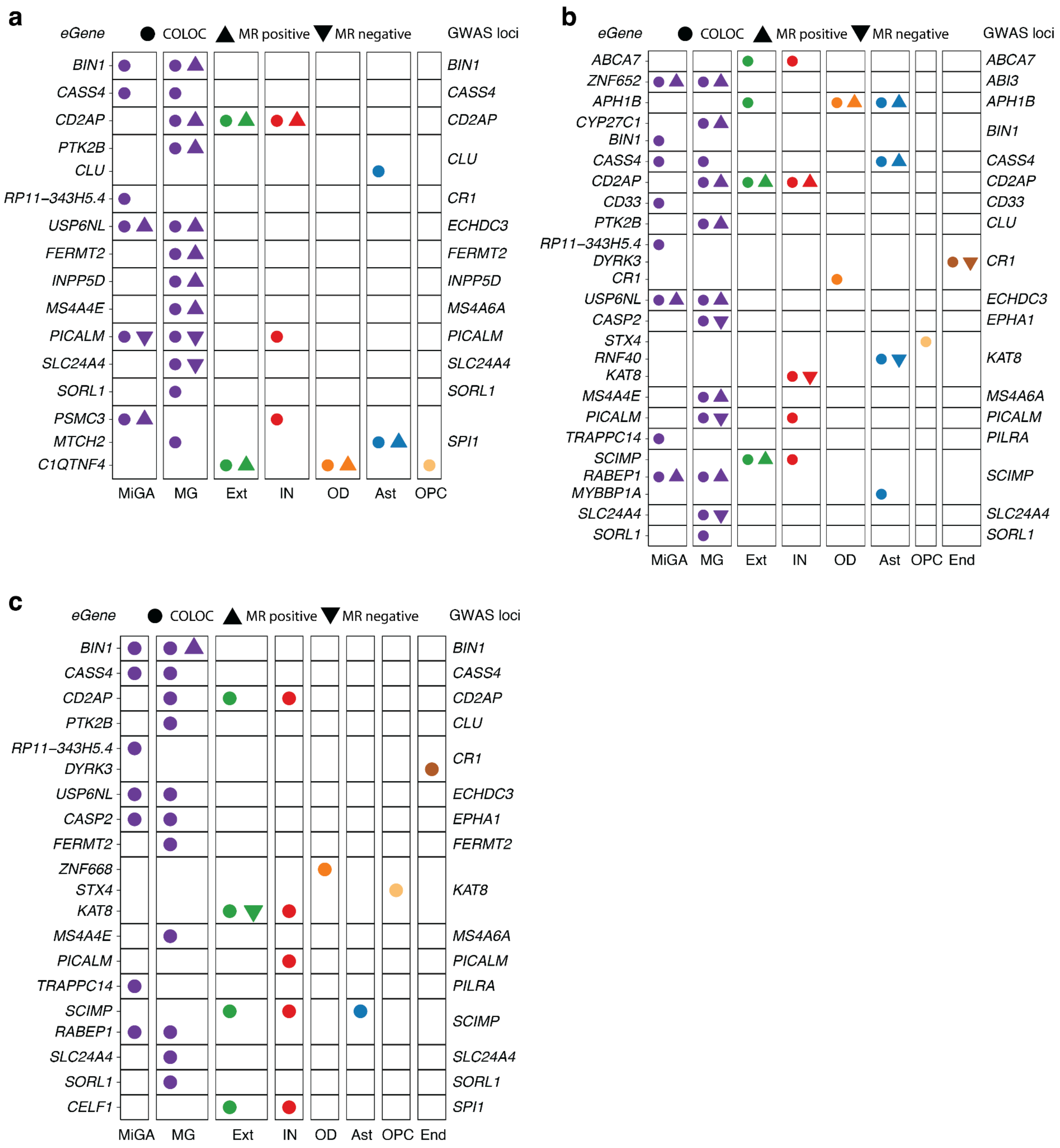


**Supplementary Figure 15 - All 3 AD GWAS loci with PP4 > 0.8 and significant MR association in SingleBrain and MiGA.**

The circle represents eQTL with PP4 > 0.8. The triangle denotes significant genes in MR results, with the orientation of the triangle corresponding to the direction of effect. **a)** *Kunkle* et al.[9] **b)** *Jansen* et al.[10] **c)** *Marioni* et al.[11] COLOC, colocalization; MR, Mendelian randomization; MiGA, Microglia Genomic Atlas; MG, microglia; Ext, excitatory neurons; IN, inhibitory neurons; Ast, astrocytes; OD, oligodendrocytes; OPC, oligodendrocyte progenitor cells; End, endothelial cells.

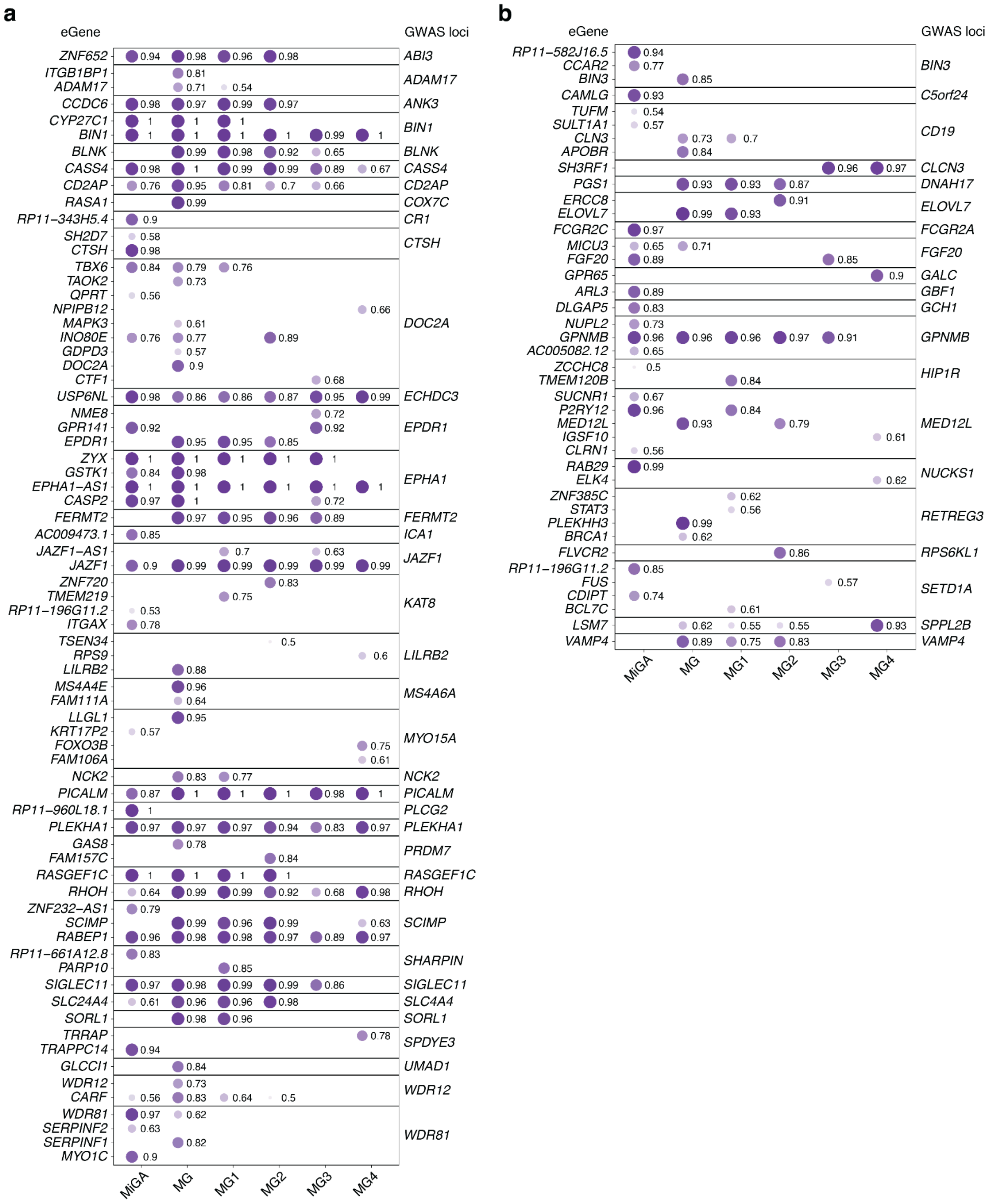

**Supplementary Figure 16 - All AD and PD GWAS locus with PP4 > 0.8 colocalization at SingleBrain microglia subtypes and MiGA.**

Shape opacity and size were scaled to the magnitude of PP4. The numbers next to the circles are PP4 values. **a)** Alzheimer's disease. **b)** Parkinson's disease. COLOC, colocalization; MiGA, Microglia Genomic Atlas; MG, microglia.

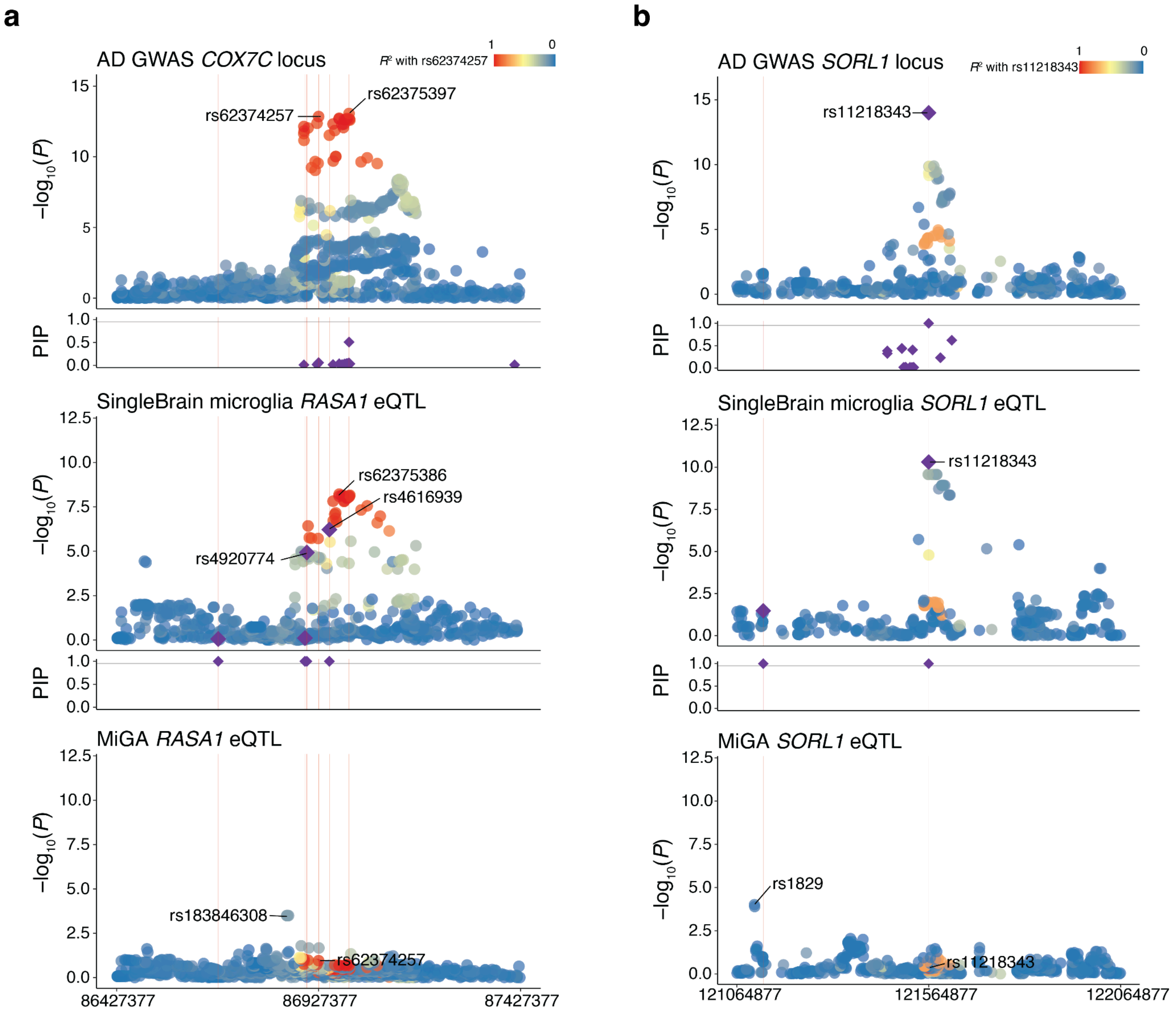


**Supplementary Figures 17 - Locus zoom and fine-mapping of *COX7C* and *SORL1* AD GWAS locus with SingleBrain MG and MiGA eQTL.**

Labels refer to lead SNPs and fine-mapping SNPs with $P < 1 \times 10^{-4}$. SNPs are colored by the LD with the lead GWAS SNP. Fine-mapping of GWAS and eQTL is present with a purple diamond under the locus zoom plot, and locus zoom only has a PIP > 0.95. **a)** The *COX7C* AD GWAS and *RASA1* eQTLs. **b)** The *SORL1* AD GWAS and *SORL1* eQTLs, respectively. AD, Alzheimer's disease; MiGA, Microglia Genomic Atlas.

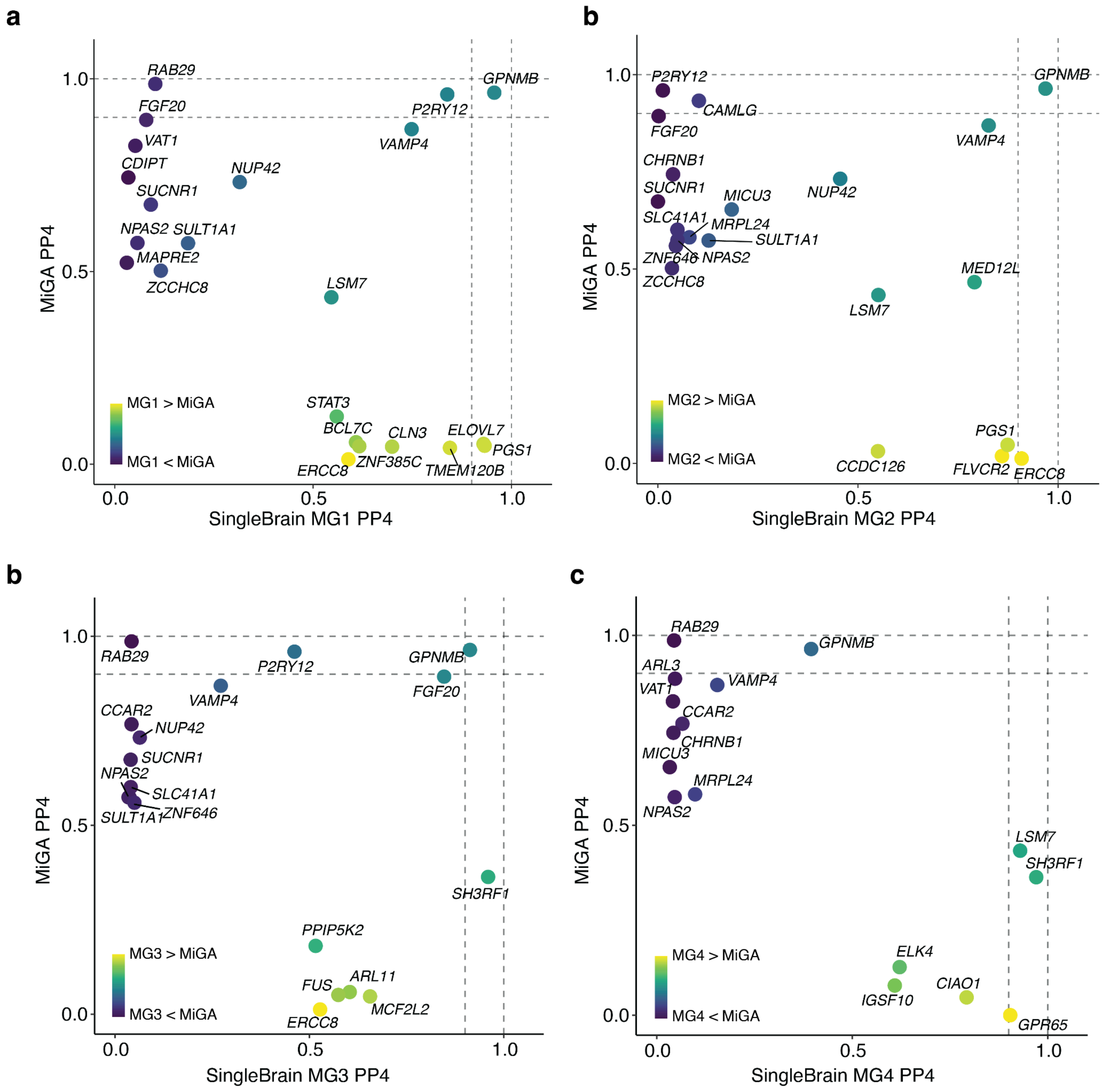


**Supplementary Figure 18 - Comparison of the maximum SingleBrain microglia subtypes and MiGA PP4 for each PD GWAS locus across both references.**

**a)** MG1 with MiGA. **b)** MG2 with MiGA. **c)** MG3 with MiGA. **c)** MG4 with MiGA, respectively.

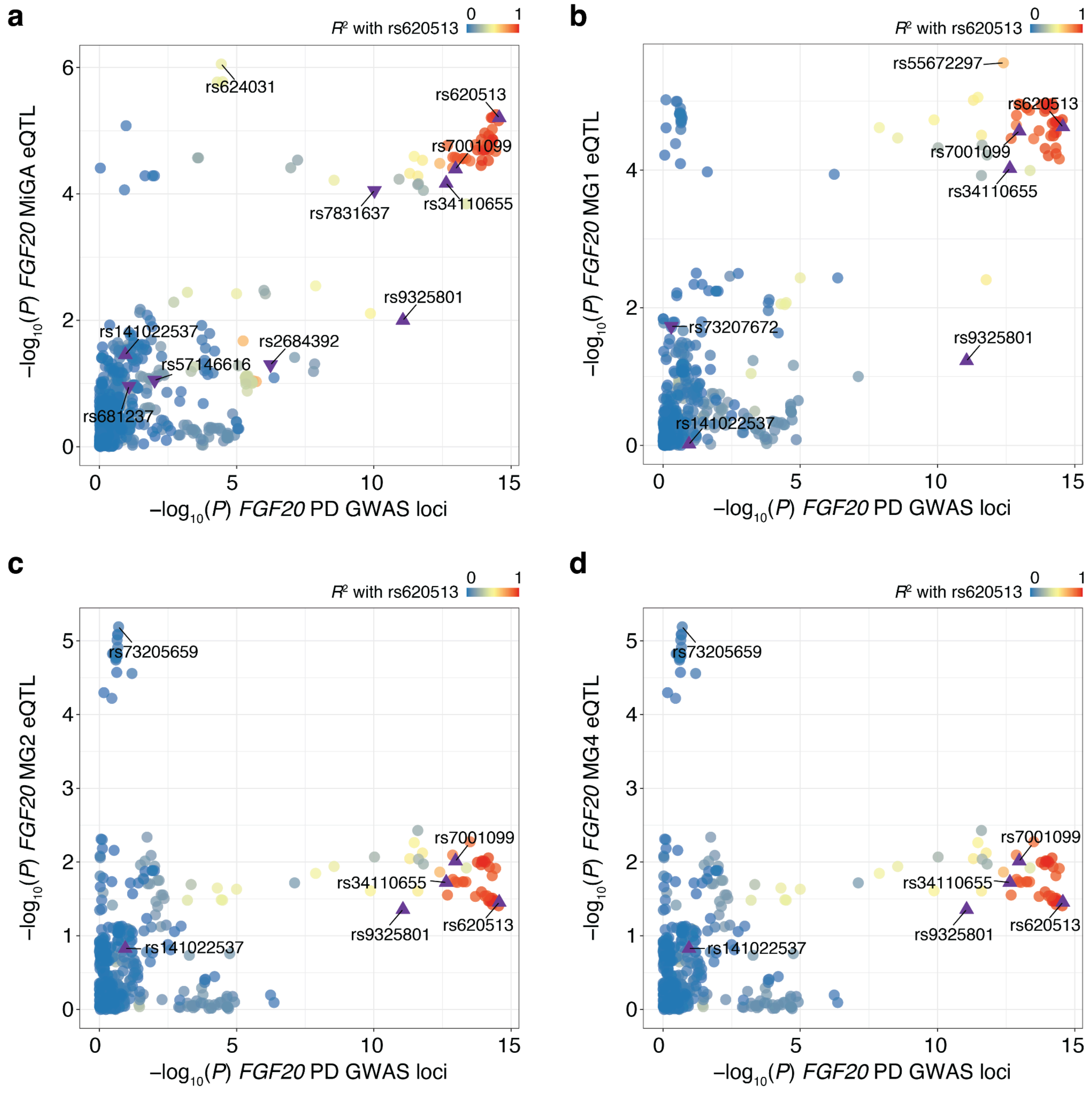


**Supplementary Figure 19 - Overlap SNPs dot plot compared to *FGF20* PD GWAS loci with *FGF20* eQTLs.**

**a)** *FGF20* PD GWAS loci with *FGF20* MiGA eQTL, **b)** *FGF20* PD GWAS loci with *FGF20* MG1 eQTL, **c)** *FGF20* PD GWAS loci with *FGF20* MG2 eQTL, **c)** *FGF20* PD GWAS loci with *FGF20* MG4 eQTL, respectively. SNPs are colored by the LD with the lead GWAS SNP. Triangles indicate GWAS fine-mapping SNPs and the inverted triangles indicate the eQTL fine-mapping SNPs (PIP > 0.95). MiGA, Microglia Genomic Atlas; MG, microglia.

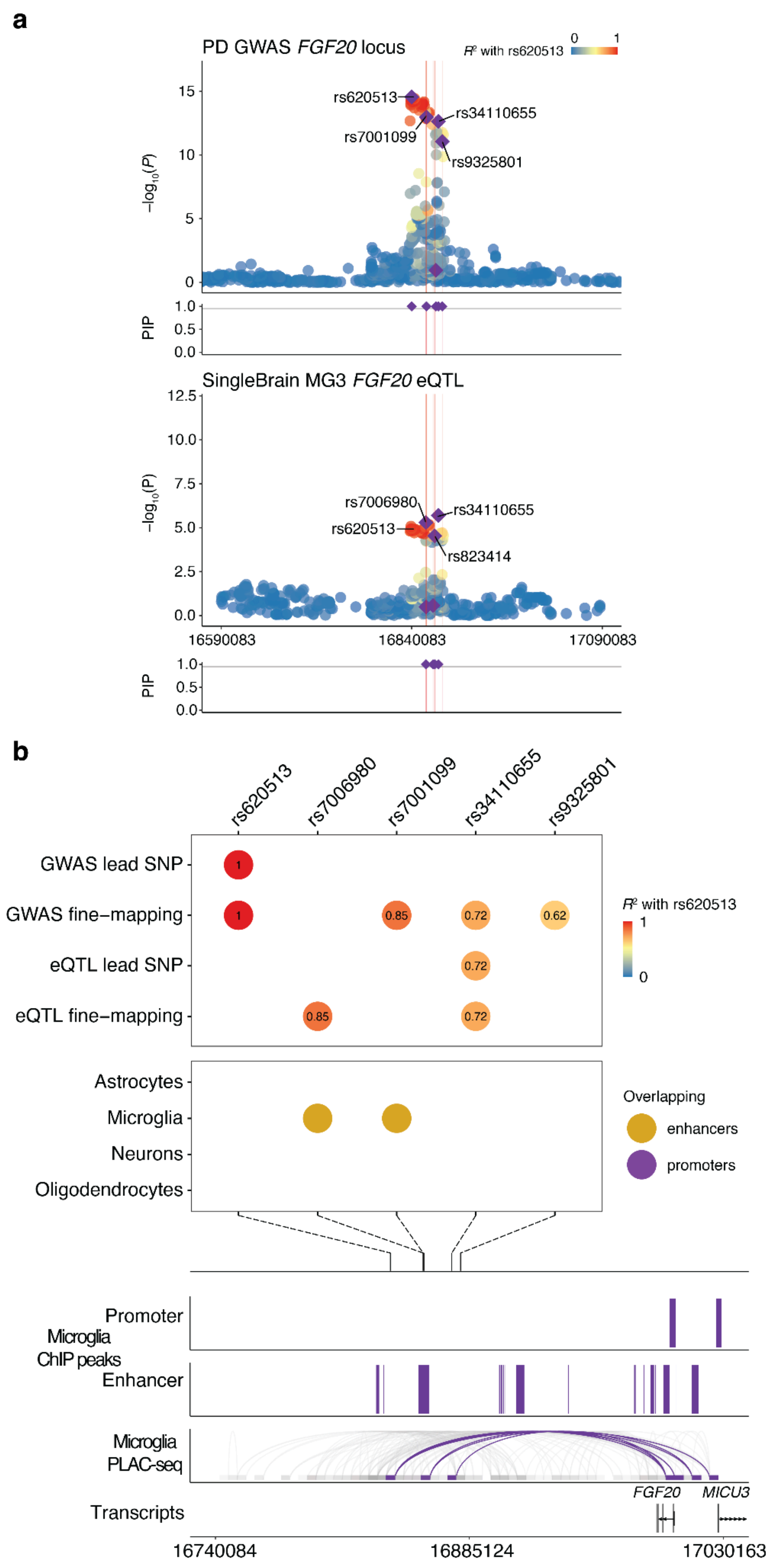


**Supplementary Figures 20 - Analysis of the *FGF20* MG3 eQTL with PD GWAS *FGF20* locus.**

**a)** Locus zoom and fine-mapping of the *FGF20* PD GWAS and *FGF20* MG3 eQTL. Labels refer to lead SNPs and fine-mapping SNPs with $P < 1 \times 10^{-4}$. SNPs are colored by the LD with the lead GWAS SNP. Fine-mapped SNPs with PIP > 0.95 from GWAS and eQTL are presented under the plot. **b)** Fine-mapping of the *FGF20* locus and combination with the GWAS lead SNP, fine-mapping SNPs, eQTL lead SNP, fine-mapping SNPs are colored by the LD with the lead GWAS SNP, overlapped with cell-type-specific enhancers and promoters were defined by *Nott* et al.[7]. Genomic plots (hg19) of the lead SNPs and fine-mapping SNPs with $P < 1 \times 10^{-4}$ and epigenomic data from the microglia ChIP-seq and PLAC-seq junctions. PD, Parkinson's disease; MG, microglia.

#### References

1. https://github.com/hail-is/hail/releases/tag/0.2.13.

2. Pedersen, B. S. *et al.* Somalier: rapid relatedness estimation for cancer and germline studies using efficient genome sketches. *Genome Med.* **12**, 62 (2020).

3. Danecek, P. *et al.* Twelve years of SAMtools and BCFtools. *Gigascience* **10**, (2021).

4. Fujita, M. *et al.* Cell subtype-specific effects of genetic variation in the Alzheimer's disease brain. *Nat. Genet.* **56**, 605–614 (2024).

5. Bryois, J. *et al.* Cell-type-specific cis-eQTLs in eight human brain cell types identify novel risk genes for psychiatric and neurological disorders. *Nat. Neurosci.* **25**, 1104–1112 (2022).

6. Humphrey, J. *et al.* Long-read RNA-seq atlas of novel microglia isoforms elucidates disease-associated genetic regulation of splicing. *medRxiv* (2023) doi:10.1101/2023.12.01.23299073.

7. Nott, A. *et al.* Brain cell type-specific enhancer-promoter interactome maps and disease-risk association. *Science* **366**, 1134–1139 (2019).

8. McAfee, J. C. *et al.* Systematic investigation of allelic regulatory activity of schizophrenia-associated common variants. *Cell Genom.* **3**, 100404 (2023).

9. Kunkle, B. W. *et al.* Genetic meta-analysis of diagnosed Alzheimer's disease identifies new risk loci and implicates Aβ, tau, immunity and lipid processing. *Nat. Genet.* **51**, 414–430 (2019).

10. Jansen, I. E. *et al.* Genome-wide meta-analysis identifies new loci and functional pathways influencing Alzheimer's disease risk. *Nat. Genet.* **51**, 404–413 (2019).

11. Marioni, R. E. *et al.* GWAS on family history of Alzheimer's disease. *Transl. Psychiatry* **8**, 99 (2018).
